# Place, Population, and Inequality: A Cross-Sectional National Analysis of Disparities in Neighborhood Physical Activity Environments Across the Urban–Rural Spectrum

**DOI:** 10.64898/2025.12.16.25342402

**Authors:** Ning Xiong, Neng Wan, Ming Wen

## Abstract

**Objectives:** Neighborhood physical activity environments (PAEs)—including walkability, recreational facilities, green space, and civic infrastructure—support active living and population health but are often inequitably distributed. This study examines racial/ethnic and socioeconomic disparities in neighborhood PAEs and assesses variation by urbanicity.

**Methods:** We used population-weighted ordinary least squares regression models with county fixed effects using the latest available 2018 data from 69,889 census tracts in the contiguous United States. Models assessed associations between racial/ethnic and poverty composition and four PAE dimensions—built, physical facilities, natural, and social environments. Sensitivity analyses compared models with and without county fixed effects and population weighting. Urbanicity-stratified models examined disparities across urban, suburban, town, rural, and mixed settings.

**Results:** Non-Hispanic White and low-poverty populations had greater access to PAEs across all domains, except that low-poverty populations lived in areas with lower walkability. Disparities were largest in urban and suburban areas. Rural high-poverty populations had more natural resources but less infrastructure and civic support. County-fixed effects reversed the walkability advantages for non-Hispanic Black observed in unadjusted models.

**Conclusions:** PAE disparities disproportionately affect racial/ethnic minority and high-poverty populations, especially in urban areas. Findings support equity-focused, context-specific interventions for environmental justice in health policy.

## 1. Introduction

Physical activity is vital for preventing chronic diseases, enhancing mental health, and promoting longevity ^1,2^. Yet, physical inactivity remains widespread in the United States (U.S.), disproportionately impacting racial and ethnic minorities and low-income populations ^3,4^. Neighborhood physical activity environments (PAEs)—like walkable streets, recreational facilities, green spaces, and social conditions—shape outdoor activity opportunities ^5^. However, access to supportive PAEs is inequitable, particularly in high-poverty and racially marginalized communities^6–8^.

These inequities reflect systemic factors, as outlined by the environmental justice (EJ) framework, which asserts that all communities deserve equal access to environmental benefits and protection from hazards, regardless of race or income ^9,10^. Historical processes like redlining have concentrated non-Hispanic Black and low-income populations in urban neighborhoods with fewer “environmental goods” and greater exposure to “environmental bads”, perpetuating health inequities through limited PAE access ^9^. This framework guides our analysis of PAE disparities, emphasizing how structural inequities shape access for marginalized communities.

Research on disparities in neighborhood PAEs reveals inconsistent findings but generally supports organizing them into four interrelated domains: built, physical facilities, natural, and social. Built environment features like walkability and transit access are generally linked to higher activity levels ^11–14^, yet paradoxically, walkability is higher in low-income or non-White neighborhoods ^15–17^. Physical facilities, such as fitness centers, are scarcer in non-Hispanic Black, Hispanic, and low-income areas ^6,18,19^, particularly in rural and disadvantaged urban neighborhoods. Natural environment, like parks, green spaces, and air quality, shapes physical activity ^20,21^. While low-income and minority communities have less park acreage ^7^, less green space access ^8,22,23^, and worse air quality ^24,25^, evidence on proximity to parks remains mixed ^7,23^. Social environment, like civic engagement and social capital, influences neighborhood capacity for physical activity ^26^. High-poverty and non-White communities often exhibit lower social capital ^27,28^, which may reduce advocacy for activity and exacerbate physical environment disparities.

Despite growing attention to PAEs, critical gaps remain. First, many studies examine only select components of PAEs, neglecting a comprehensive, multidimensional assessment. Second, most focus on urban areas, limiting relevance across the urban–rural spectrum. Third, prior work conflates race/ethnicity and socioeconomic status (SES) effects, obscuring whether PAE disparities stem from race, SES, or their intersection. Methodological limitations, including reliance on unweighted models and failure to address higher geographic-scale confounding, undermine generalizability and contribute to inconsistent findings. These gaps underscore the need for a comprehensive, integrated approach to studying national PAE disparities.

This study addresses these gaps by applying the Neighborhood Environment Framework for Outdoor Physical Activity, a literature-informed structure we developed to organize PAEs into four domains: built, physical facilities, natural, and social environments (Figure 1) ^6–8,11–28^. Using 69,889 U.S. census tracts, we examine associations between neighborhood race/ethnicity and poverty levels and PAEs across the urban–rural continuum. We use models with and without county fixed effects and population weighting to distinguish national vs. within-county disparities and place-vs. population-based effects, and further test models including and excluding race/ethnicity or poverty to assess their independent contributions. Grounded in the EJ framework, we hypothesize that non-Hispanic Black, Hispanic, and low-income populations have less access to supportive PAEs, particularly in urban areas.

**Figure 1.**
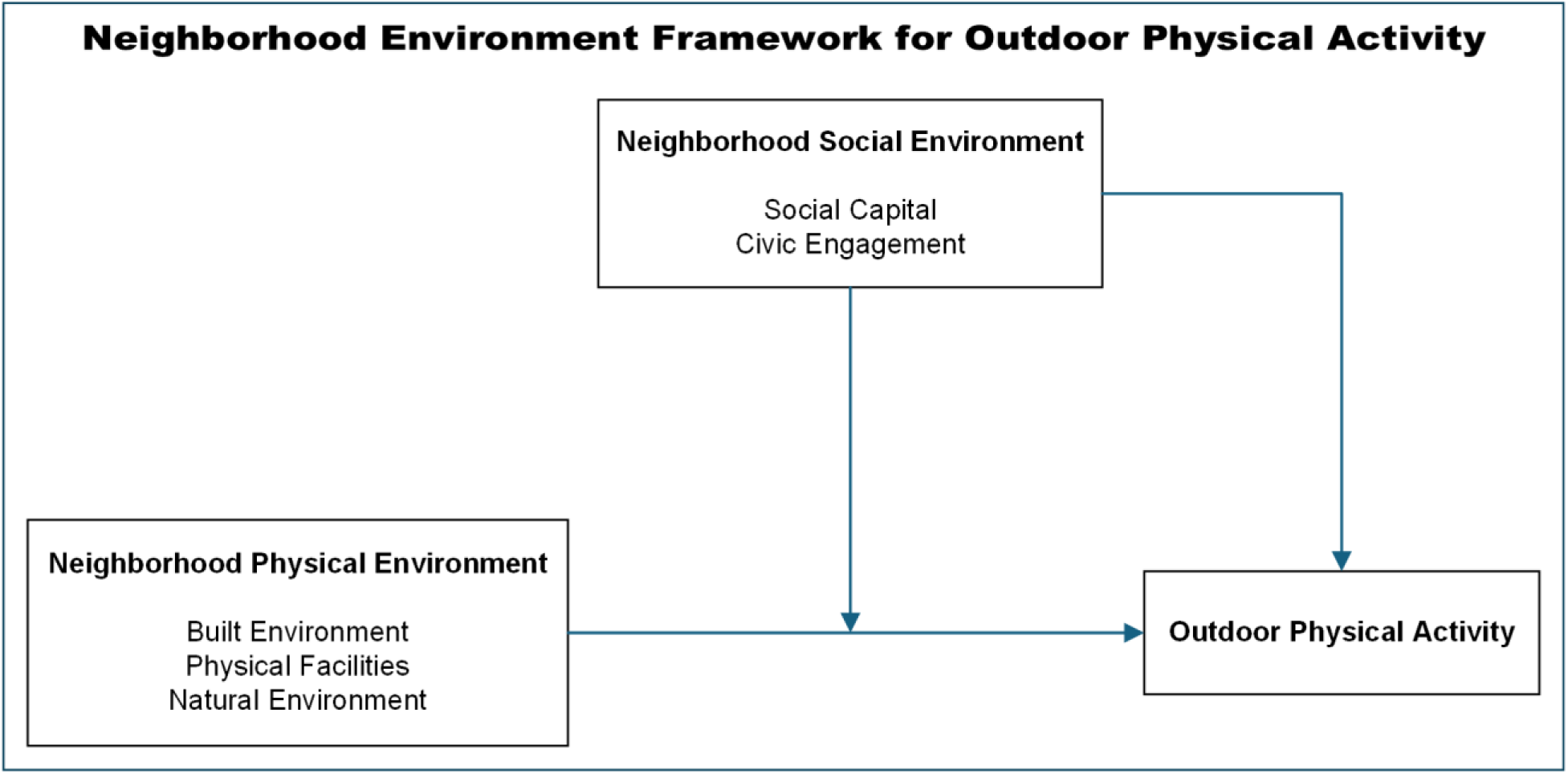
Neighborhood Environment Framework for Outdoor Physical Activity. This figure depicts a literature-informed framework used to guide the measurement and analysis of neighborhood physical activity environments (PAEs). The framework encompasses four domains—built, physical facilities, natural, and social environments—and illustrates pathways through which neighborhoods shape outdoor physical activity. In this study, the framework provides a descriptive structure for examining disparities in PAEs.

## 2. Methods

### 2.1 Study Area and Sample

The study area includes census tracts in the contiguous U.S. We excluded Alaska, Hawaii, and U.S. Territories due to their distinct geographic, environmental, and urban development characteristics, which could bias or limit generalizability. From 72,538 census tracts defined by 2010 boundaries, we excluded 609 zero-population tracts and 12 with invalid urbanicity codes, then removed 2,028 tracts with missing PAE data, yielding a final sample of 69,889 tracts—96.35% of all census tracts, which covers 97.65% of the 2018 population in the contiguous U.S.

### 2.2 Data and Measures

We used census tract-level data from the 2014–2018 American Community Survey (ACS), supplemented with multiple national sources, to examine disparities in neighborhood PAEs by racial/ethnic and SES composition across levels of urbanicity. All data were from 2018 or the nearest available year to ensure temporal consistency across sources, as 2018 is the most recent year available for national built environment indicators in the Smart Location Database (SLD) (Version 3.0). Population counts from the ACS were used to construct population-weighted models to reflect residents’ exposure to local environments.

Neighborhood sociodemographic composition was defined using race/ethnicity and poverty indicators. Racial/ethnic composition, sourced from ACS, was categorized into five mutually exclusive groups: non-Hispanic White (>50%), non-Hispanic Black (>50%), Hispanic/Latino (>50%), non-Hispanic Asian (>50%), and mixed (no group >50%). SES composition, based on tract-level individual poverty rate from ACS, was classified as low (<10%), medium (10–20%), or high (>20%). Urbanicity from the U.S. Environmental Protection Agency was included as a covariate and used to stratify models. It was measured using the National Center for Education Statistics urban–rural locale classification (1–5 scale), capturing varying levels of development and infrastructure across tracts and more closely reflecting the rural/urban nature of the immediate environment.

Dependent variables spanned two broad PAE dimensions: physical (built, facilities, natural) and social environment. Built environment indicators derived from SLD (land use mix, road network/street intersection density, transit access). Physical facilities, sourced from the National Neighborhood Data Archive (NaNDA) ^29^, included counts of recreational and physical fitness centers, membership sports and recreation clubs, specialized recreational establishments, and public golf courses and driving ranges. Natural environment indicators reflected access to green space and exposure to environmental hazards. These included park cover (percent of recreationally accessible parks), summertime maximum values of the Normalized Difference Vegetation Index (NDVI), both sourced from the Open Science Framework ^30^, green space rate (2019 National Land Cover Database), and tree canopy cover (NLCD 2018 USFS Dataset). We also included average annual concentrations of PM2.5 (µg/m³) as an air pollution measure, obtained from the Center for Air, Climate, and Energy Solutions.

As a social environment indicator, we used the 2020 Census self-response rate as a proxy for neighborhood-level civic engagement and social capital, consistent with prior research ^31^. This measure complements physical and natural environment indicators by capturing trust, participation, and responsiveness. To align 2020 data with 2010 census tract boundaries, we weighted each 2020 tract’s self-response rate by the proportion of its land area overlapping with corresponding 2010 tracts, using the U.S. Census Bureau’s 2020 Tract Relationship Files, ensuring consistent spatial alignment for regression modeling. Variable definitions are summarized in Table 1, with 2010 census tract boundaries used throughout.

**Table 1.**
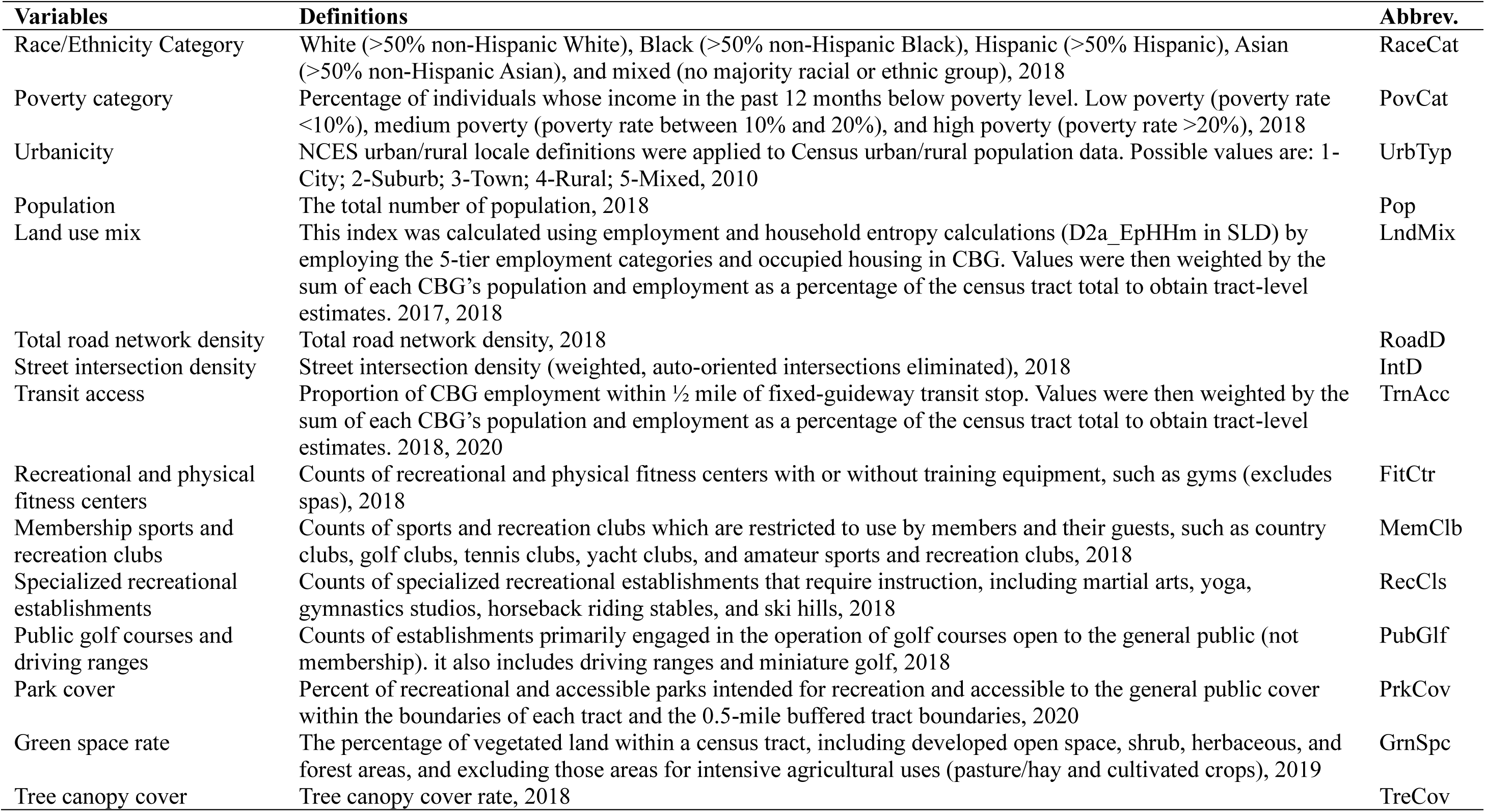
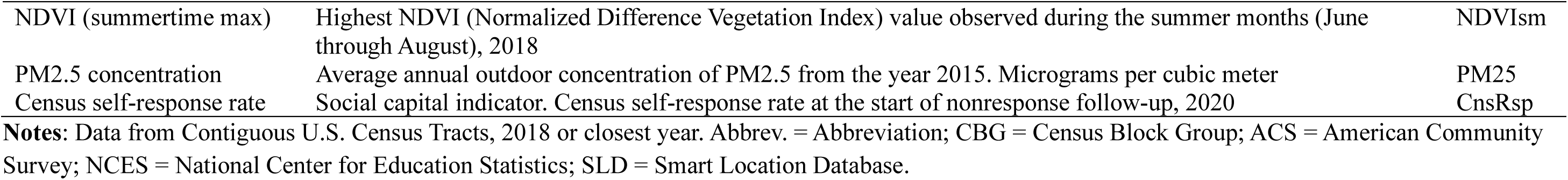
Variables and Definitions Used in the Analysis of Neighborhood Physical Activity Environments, Contiguous United States Census Tracts, 2018.

### 2.3 Analytical Strategies

Ordinary least squares (OLS) regression models assessed relationships between dependent variables (physical and social environment) and independent variables (neighborhood poverty, racial/ethnic composition), with urbanicity as a covariate. County-fixed effects and population weights were included to account for unobserved, time-invariant county-level factors and isolate within-county disparities, which may better approximate conditions experienced by residents rather than treating all tracts equally regardless of population size.

We conducted several sensitivity analyses to assess robustness. First, separate models estimated the independent effects of poverty or racial/ethnic composition without urbanicity as a covariate. Second, models without county-fixed effects captured national-level disparities, distinguishing national from within-county disparities. Third, unweighted models complement population-weighted estimates to assess geographic versus population-driven disparities, offering a nuanced view of environmental inequities while recognizing the ecological nature of the data.

Models were stratified by urbanicity (urban, suburban, town, rural, mixed) to explore contextual variations, retaining all racial/ethnic categories (including smaller groups like Asians), though coefficients for limited-sample categories were interpreted cautiously. PAE variables were standardized (mean = 0, SD = 1) to enable effect size comparisons across outcomes.

## 3. Results

### 3.1 Descriptive Statistics

Table 2 summarizes the descriptive statistics for PAE by urbanicity. Urban tracts showed the highest infrastructure levels but the lowest green infrastructure. Rural tracts had minimal built environment features, but the highest natural features. Suburban areas displayed moderate infrastructure and green space. Towns exhibited a relatively high land use mix but the lowest park coverage, while mixed areas had intermediate values. Fitness center followed an urban-rural gradient: 1.07 in urban, 1.18 in suburban, and 0.23 in rural tracts. Social environment measured by census self-response rate was highest in suburban areas and lowest in rural tracts. Supplemental materials (Table A1, Figures A1–A3) detail tract distribution and spatial patterns by race/ethnicity, poverty, and urbanicity across 69,889 U.S. tracts, with further analysis in the Supplemental “Descriptive Statistics (Detail)” section. These patterns highlight geographic variation in PAEs, examined further in the OLS regression analyses below.

**Table 2.** Mean Physical Activity Environment Indicators by Urbanicity, Contiguous United States Census Tracts, 2018.

| Indicators | National | Urban | Suburban | Town | Rural | Mixed |
| --- | --- | --- | --- | --- | --- | --- |
| LndMix | 0.524 | 0.510 | 0.530 | 0.610 | 0.480 | 0.550 |
| RoadD | 14.451 | 22.990 | 17.000 | 12.620 | 2.340 | 5.790 |
| IntD | 69.405 | 120.160 | 80.960 | 66.030 | 2.850 | 17.800 |
| TrnAcc | 0.066 | 0.160 | 0.050 | 0.000 | 0.000 | 0.000 |
| FitCtr | 0.942 | 1.070 | 1.180 | 0.970 | 0.230 | 0.860 |
| MemClb | 0.605 | 0.590 | 0.660 | 0.570 | 0.400 | 0.700 |
| RecCls | 0.380 | 0.430 | 0.520 | 0.320 | 0.070 | 0.310 |
| PubGlF | 0.117 | 0.050 | 0.090 | 0.100 | 0.160 | 0.230 |
| PrkCov | 0.074 | 0.060 | 0.070 | 0.050 | 0.100 | 0.090 |
| GrnSpc | 0.297 | 0.140 | 0.250 | 0.290 | 0.530 | 0.460 |
| TreCov | 0.223 | 0.130 | 0.210 | 0.160 | 0.360 | 0.310 |
| NDVIsm | 0.617 | 0.490 | 0.590 | 0.640 | 0.790 | 0.730 |
| PM25 | 8.055 | 8.730 | 8.310 | 7.900 | 6.850 | 7.460 |
| CnsRsp | 0.626 | 0.600 | 0.690 | 0.600 | 0.540 | 0.640 |

**Notes:** This table presents the mean values of physical activity environment indicators across census tracts stratified by urbanicity: urban, suburban, town, rural, and mixed areas, as well as national averages. Data from Contiguous U.S. Census Tracts, 2018 or closest year.

### 3.2 Results of OLS Regressions

#### 3.2.1 National-Level Results

Table 3 shows racial/ethnic and socioeconomic differences in access to built, physical facility, natural, and social environment dimensions of PAE. Non-Hispanic Black and Hispanic groups show lower access to favorable built environment features, physical facilities, green spaces, and civic engagement, with higher air pollution exposure, while medium- and high-poverty populations have higher walkability indicators. These findings, from population-weighted models with county-fixed effects (Model 1, Table 3), may better reflect residents’ lived conditions rather than tract characteristics due to the weighting. Non-Hispanic Black and Hispanic people had significantly lower scores on desirable built environment features like land use mix (B = −0.534, −0.305) and street intersection density (B = −0.065, 0.011), while medium- and high-poverty people had higher walkability (B = 0.078, 0.152 for land use mix; B = 0.181, 0.363 for intersection density). These disadvantaged populations also experienced significantly lower availability of physical fitness centers (e.g., B = −0.515 for non-Hispanic Black; B = −0.604 for Hispanic; B = −0.147 for high poverty), green infrastructure like tree canopy cover (B = −0.100, −0.208, and −0.181, respectively) and park coverage (−0.080, −0.260, and −0.067, respectively), and census self-response rate (B = - 0.508, −0.440, and −0.972, respectively), with higher PM2.5 exposure (Black: B = 0.177; Hispanic: B = 0.378; medium/high poverty: B = 0.106, 0.178). Disparities in the social environment were especially pronounced, with the largest gap in the census self-response rate observed among people in high poverty areas (B = −0.972).

**Table 3.** Associations Between Neighborhood Sociodemographic Composition and Physical Activity Environment Indicators, Contiguous United States Census Tracts, 2018.

| Indicators | B_NH >50% | Hisp >50% | A_NH >50% | MedPov | HighPov |
| --- | --- | --- | --- | --- | --- |
| LndMix | -0.534** | -0.305** | -0.008 | 0.078** | 0.152** |
| RoadD | -0.013** | 0.084** | 0.031 | 0.157** | 0.292** |
| IntD | -0.065** | 0.011 | -0.068** | 0.181** | 0.363** |
| TrnAcc | -0.154** | -0.013 | -0.113** | 0.100** | 0.223** |
| FitCtr | -0.515** | -0.604** | -0.383** | -0.064** | -0.147** |
| MemClb | -0.390** | -0.394** | -0.250** | -0.099** | -0.133** |
| RecCls | -0.462** | -0.467** | -0.158** | -0.050** | -0.100** |
| PubGlF | -0.077** | -0.118** | 0.025 | -0.137** | -0.209** |
| PrkCov | -0.080** | -0.268** | -0.101** | -0.063** | -0.067** |
| GrnSpc | -0.076** | -0.329** | -0.180** | -0.147** | -0.228** |
| TreCov | -0.100** | -0.208** | -0.173** | -0.088** | -0.181** |
| NDVIsM | 0.037** | -0.198** | -0.159** | -0.124** | -0.241** |
| PM25 | 0.177** | 0.378** | 0.258** | 0.106** | 0.178** |
| CnsRsp | -0.508** | -0.440** | -0.079** | -0.460** | -0.972** |

**Note:** Estimates are standardized regression coefficients from multivariable linear regression, including race/ethnicity, poverty, which are weighted by population with urbanicity and county fixed effects. Statistical significance is denoted as follows: \*\* $p < 0.05$ ; \* $p < 0.10$ . Reference categories are the majority non-Hispanic White (race) and low poverty (<10%) (poverty). Race/ethnicity categories include: Non-Hispanic White (W\_NH), non-Hispanic Black (B\_NH), Hispanic (Hisp), non-Hispanic Asian (A\_NH), and Mixed (no majority racial or ethnic group). Poverty categories are defined as: Low poverty (<10%) (LowPov), medium poverty (10–20%) (MedPov), and high poverty (>20%) (HighPov). All variables are from 2018 or the closest available year.

#### 3.2.2 Sensitivity Analyses

Supplemental Table A2 shows that PAE disparities persist across model specifications. Non-Hispanic Black, Hispanic, and high-poverty populations show lower access to fitness centers, green spaces, and civic engagement, with higher air pollution exposure, though walkability varies by county-fixed effects. These robustness checks are further detailed in the analyses below, and full results are provided in the Sensitivity Analyses (Detail) section of the Supplemental Materials.

First, we estimated three models to assess the independent and joint effects of racial/ethnic composition and poverty on neighborhood PAEs. models estimating race/ethnicity or poverty effects independently (Supplemental Table A2, Models 1–2) showed consistent disadvantages across most PAE domains, even where walkability indicators appeared higher. In fully adjusted models including both race/ethnicity and poverty (Table 3, Model 1), disparities remained largely significant, though some associations attenuated.

Second, to evaluate the robustness of disparities in PAEs, we compared four model specifications (Table 3, Supplemental Table A3)—population-weighted with county fixed effects, unweighted with county fixed effects, population-weighted without county fixed effects, and unweighted without county fixed effects. Without county-fixed effects, non-Hispanic Black people/tracts showed greater walkability (e.g., road network density, street intersection density, and transit access), but this reversed after controlling for county, suggesting within-county inequities. Other disadvantages remained consistent. For Hispanic people/tracts, some built and natural environment disparities weakened with county-fixed effects. For high-poverty people/tracts, some walkability indicators increased with fixed effects, while deficits in the social environment and some natural indicators persisted or reversed.

Third, comparing weighted and unweighted models revealed that weighting slightly shifted coefficient magnitudes but did not alter key patterns. Disparities in built, natural, and social environments for non-Hispanic Black, Hispanic, and high-poverty populations remained consistent, reinforcing robustness across specifications.

#### 3.2.3 Urbanicity-Stratified Results

Supplemental Table A4 presents urbanicity-stratified model results testing PAE disparity persistence, showing that non-Hispanic White, low-poverty populations have greater access across favorable built, physical facilities, natural environment, and social environment in urban, suburban, and rural settings, with larger gaps in urban and suburban areas. Non-Hispanic Black and Hispanic populations exhibit lower access to some built environment features in urban and suburban contexts, while high-poverty populations show higher walkability in towns; all experience reduced access to recreational facilities and civic engagement, alongside higher air pollution exposure. These patterns, detailed below, stem from Model 1 (population-weighted, county-fixed effects, including race/ethnicity and poverty) in Supplemental Table A3. The remaining specifications (Supplemental Tables A5–A7) show similar patterns.

Stratified analyses showed that the magnitude and direction of disparities varied substantially by urbanicity. In urban areas, disparities in built environment access remained significant and often intensified. For example, non-Hispanic Black and Hispanic populations had lower land use mix (B = −0.630, −0.397) and fitness center access (B = −0.539, −0.714). Suburban areas showed similar gaps (e.g., street intersection density: B = −0.060, 0.084; fitness centers: B = - 0.588, −0.340). In town areas, associations were less consistent but notable; high-poverty people had higher PM2.5 (B = 0.132) and lower census self-response rate (B = −0.970), while Hispanic people experienced higher land use mix (B = 0.493). In rural areas, high-poverty people had significantly higher tree canopy cover (B = 0.315) and PM2.5 (B = 0.098) but lower census self-response rate (B = −0.377); however, built environment features like transit access and walkability were generally weaker or non-significant. In mixed areas, disparities persisted across PAE domains, particularly for green infrastructure and civic engagement, with non-Hispanic Black and Hispanic people experiencing lower tree canopy cover (B = −0.135 and −0.287) and census self-response rate (B = - 0.361 and −0.439).

## 4. Discussion

This study provides a comprehensive lens on how place, population, and inequality intersect in their association with PAEs across the urban–rural spectrum in the contiguous U.S., extending the application of the EJ framework ^9,10^. The Neighborhood Environment Framework for Outdoor Physical Activity, a literature-informed organizing structure, groups built environment, physical facilities, natural environment, and social environment indicators to guide our analysis. We examine associations between neighborhood race/ethnicity and poverty levels and PAEs across the urban–rural continuum. Dual-modeling approaches capture disparities: population-weighted OLS regressions highlight exposure-based inequities experienced by non-Hispanic Black, Hispanic, and poverty urban residents, while unweighted models emphasize tract-level disparities. Results confirm significant PAE disparities, especially in urban areas, with context-specific patterns informing equity-focused interventions.

Grounded in the EJ framework, historical practices like segregation and redlining have concentrated marginalized groups in areas with fewer “environmental goods” (e.g., parks) and higher “environmental bads” (e.g., pollution) ^9,10^. Using OLS models with county-fixed effects and population weights, this study shows systemic disadvantages for non-Hispanic Black, Hispanic, and high-poverty populations across PAE dimensions, consistent with prior evidence ^16,24^. In contrast, non-Hispanic White and low-poverty populations exhibit systemic privilege ^9^, with greater access to favorable physical facilities ^18^, natural environment ^8^, and social environment ^27,28^. Although non-Hispanic White populations have higher access to walkability, this contrasts with prior studies reporting greater walkability in non-White neighborhoods ^15–17^. High-poverty groups also show greater walkability, challenging the universal wealth advantage. These patterns reveal that built environment advantages do not consistently align with race or SES, underscoring the need for equity-focused strategies.

Methodologically, this study improves on prior work through diverse model specifications. A key finding is the reversal of walkability results with and without county-fixed effects. Without them, non-Hispanic Black populations show greater walkability—reflected in road network density, street intersections, and transit access—but these advantages reverse with county-fixed effects. This suggests that while non-Hispanic Black people may reside in walkable counties, they face disadvantages within those counties. Comparing weighted and unweighted models shows minimal changes in effect direction or significance, confirming the robustness of findings. These results support both population- and place-based interpretations, helping target interventions based on who is most affected and where disparities are concentrated.

Urbanicity-stratified analyses reveal diverse PAE disparities, with the most pronounced disparity in urban and suburban areas. In urban and suburban areas, White and low-poverty groups enjoy greater access to facility access ^6^ and green infrastructure ^12^. Rural high-poverty populations have more natural resources but lack infrastructure and civic support. In town and mixed areas, non-Hispanic White populations retain social strengths, but high-poverty populations face air quality issues. Rural/urban disparities are often related more to poverty than race, shown by sparse infrastructure. High-poverty populations in rural areas are more advantaged in natural environment indicators. The absence of a universal White and wealth advantage in rural settings suggests geographic moderation of historical inequities, supporting tailored interventions across urbanicity.

This study extends the EJ framework by disentangling the association of race/ethnicity and poverty with PAEs ^16^. Though disparities attenuate in combined models, persistent disadvantages for non-Hispanic Black and Hispanic groups suggest distinct and cumulative effects. These findings reinforce that EJ stems from structural racism, not just economic deprivation ^9,32^. While high-poverty groups may benefit from built environment features, they remain disadvantaged in facilities, natural quality, and social engagement. The persistence of disparities across model specifications highlights the systemic nature of these inequities.

## 5. Limitations

This study is subject to several limitations. First, its cross-sectional design limits causal inference and prevents evaluation of temporal trends. Second, neighborhood sociodemographic composition was based on majority-group thresholds, which may obscure intra-tract diversity. Third, PAEs were measured using objective indicators and did not capture perceived safety, access, or quality—factors known to influence physical activity. Finally, while county-fixed effects help isolate within-county disparities, unmeasured confounding at more localized scales may remain. Future research should incorporate longitudinal and mixed-methods approaches to better understand temporal dynamics and community experiences of neighborhood environments.

## 6. Conclusions

Non-Hispanic White and low-poverty populations generally have greater access to neighborhood PAEs across multiple domains. However, low-poverty populations lived in areas with lower walkability, highlighting variation across PAE components. Disparities were most pronounced in urban and suburban areas, where non-Hispanic Black, Hispanic, and high-poverty populations experienced consistent disadvantages. In rural areas, high-poverty populations had relatively greater access to natural resources but lacked supporting infrastructure and civic engagement opportunities. Adjusting for county-fixed effects reversed the apparent walkability advantage observed for non-Hispanic Black populations in unadjusted models, indicating that spatially aggregated data may obscure localized inequities. These findings emphasize the need for equity-focused, context-sensitive strategies to address neighborhood PAE disparities and promote physical activity across diverse geographic settings.

## Supporting information

PAE18_Supplemental Materials_20250830

## Data Availability

This study was conducted using publicly available, de-identified secondary data and was exempt from institutional review board oversight.

## Acknowledgements

This work was supported by the National Cancer Institute of the National Institutes of Health under award number R37CA276365. The content is solely the responsibility of the authors and does not necessarily represent the official views of the National Institutes of Health.

