## Supplementary material for "Place, Population, and Inequality: A Cross-Sectional National Analysis of Disparities in Neighborhood Physical Activity Environments Across the Urban–Rural Spectrum": PAE18_Supplemental Materials_20250830

### 1. Sensitivity Analyses (Detail)

#### 1.1 Descriptive Statistics

Supplemental Table A1 presents the distribution of tracts by racial/ethnic composition and poverty level across urbanicity types. Percentages reported in the text are based on the full sample, while Supplemental Table A1 provides corresponding tract counts. Nationally, 69.0% of tracts were majority non-Hispanic White, 8.2% were majority non-Hispanic Black, 9.4% were majority Hispanic or Latino, 0.8% were majority non-Hispanic Asian, and 12.5% were classified as racially/ethnically mixed (no group >50%). In terms of socioeconomic composition, 41.8% of tracts were categorized as low-poverty, 32.3% as medium-poverty, and 25.9% as high-poverty. Tracts were distributed across urbanicity as follows: 31.3% urban, 30.6% suburban, 13.9% mixed, 13.9% rural, and 4.9% town.

Supplemental Figures A1–A3 illustrate the spatial distributions of racial/ethnic composition, poverty levels, and urbanicity across 69,889 census tracts in the contiguous U.S., highlighting broad patterns rather than precise local variations due to the national scale. Supplemental Figure A1 shows majority non-Hispanic Black populations concentrated in the Southeast, urban Midwest, and Northeast, majority Hispanic populations along the Southwest and in select urban centers, and majority non-Hispanic White populations dominating the Midwest and Mountain West. Supplemental Figure A2 maps low poverty in suburban rings and affluent rural areas, medium poverty across semi-urban and transitional zones, and high poverty in the rural South, urban cores, and border regions. Supplemental Figure A3 depicts urban tracts clustered in major metropolitan areas, suburban rings encircling these cores, towns scattered as hubs in less dense regions, rural expanses covering the Great Plains and Mountain West, and mixed areas in peri-urban transitions. Together, these maps underscore the geographic heterogeneity of racial/ethnic, socioeconomic, and urbanicity factors influencing PAE disparities.

#### 1.2 Comparison of Independent and Joint Effects

We estimated three models to examine the independent effects of racial/ethnic composition and poverty on PAEs and assess whether disparities persist when both factors are considered jointly. These included a model with racial/ethnic composition alone (Model 2 of Table 3), one with poverty alone (Model 3 of Table 3), and one with both and urbanicity included as a covariate (Model 1 of Table 3).

In the race/ethnicity-only model (Model 2 of Table 3), non-Hispanic Black and Hispanic people were disadvantaged across most PAE outcomes compared to non-Hispanic White people. Specifically, they had significantly lower scores for walkability indicators such as land use mix (B = -0.421 and -0.192, respectively), and physical facility access, including recreational and physical fitness centers (B = -0.532 and -0.640, respectively). They also faced notably lower levels of green infrastructure, such as tree canopy cover (B = -0.252 and -0.358), park coverage (B = -0.148 and -0.339), green space coverage (B = -0.314 and -0.559), and NDVI (B = -0.161 and -0.395), and markedly lower civic engagement indicated by census self-response rates (B = -0.997 and -0.974). Conversely, non-Hispanic Black and Hispanic people exhibited higher street intersection density (B = 0.288 and 0.349) and road network density (B = 0.320 and 0.397). Additionally, they experienced significantly higher exposure to PM2.5 air pollution (B = 0.336 and 0.535).

In the poverty-only model (Model 3 of Table 3), associations were mixed. medium- and high-poverty people had significantly higher street intersection density (B = 0.272 and 0.586), and road network density (B = 0.267 and 0.562). However, they also exhibited significantly reduced access to physical facilities like recreational and physical fitness centers (B = -0.122 and -0.276), and green infrastructure including lower park access (B = -0.115 and -0.192), green spaces (B = -0.255 and -0.486), and tree canopy coverage (B = -0.152 and -0.336), and NDVI (B = -0.189 and -0.390). These people also saw substantially lower civic engagement, as reflected by census self-response rates (B = -0.545 and -1.158), and elevated PM2.5 concentrations (B = 0.186 and 0.379).

When racial/ethnic composition and poverty were simultaneously included along with urbanicity (Model 1 of Table 3), most associations attenuated slightly but remained significant, suggesting overlapping yet distinct impacts. In this combined model, disparities persisted with non-Hispanic Black and Hispanic people consistently experiencing lower access to recreational and physical fitness centers (B = -0.515 and -0.604), reduced green infrastructure such as tree canopy coverage (B = -0.100 and -0.208), and substantially lower census self-response rates (B = -0.508 and -0.440). Although street intersection density and road network density advantages initially observed for non-Hispanic Black and Hispanic people in race/ethnicity-only models reversed or greatly weakened (e.g., road network density shifted to B = -0.013 for non-Hispanic Black people and B = 0.084 for Hispanic people), PM2.5 exposure remained disproportionately high (B = 0.177 and 0.378). Likewise, high-poverty people maintained higher intersection density (B = 0.363) and road network density (B = 0.292), but their disadvantages in physical facilities (B = -0.147), less green space (e.g., tree canopy cover B = -0.181), and civic engagement (B = -0.972) continued to be significant and substantial.

To explore how these disparities vary by geographic context, we further examined the independent and joint effects of racial/ethnic composition and poverty within each urbanicity category—urban, suburban, town, rural, and mixed. Stratified results (Supplemental Tables A3–A6) indicate that both race/ethnicity and poverty independently contribute to PAE disparities across urbanicity types.

#### 1.3 Comparison Across Model Specifications

To assess the robustness of observed disparities and their sensitivity to modeling choices, we compared estimates from four specifications: 1) population-weighted with county fixed effects (Table 3); 2) unweighted with county fixed effects (Supplemental Table A2); 3) population-weighted without county fixed effects (Supplemental Table A2); and 4) unweighted without county fixed effects (Supplemental Table A2). The analysis is organized into two subsections: 1) comparing models with and without county-fixed effects; and 2) comparing models with and without population weighting, focusing on national-level results from Model 1.

##### 1.3.1 With and Without County Fixed Effects

To capture different dimensions of disparity, we estimated two sets of models: one without county-fixed effects to capture national-level disparities across all tracts, and another with county-fixed effects to examine whether racial/ethnic and socioeconomic disparities persist within the same counties, allowing us to isolate disparities within counties. In Model 1, the inclusion of county fixed effects significantly altered the magnitude and direction of associations, highlighting their sensitivity to geographic controls. For the non-Hispanic Black people (in weighted models) or tracts (in unweighted models) (relative to the majority non-Hispanic White), built environment indicators showed striking changes. In models without county fixed effects, non-Hispanic Black people/tracts appeared to have greater walkability infrastructure access, with weighted estimates showing positive associations for road network density (B = 0.141), intersection density (B = 0.039), and transit access (B = 0.149). The unweighted estimates were similarly positive (road network density, B = 0.151; intersection density,

B = 0.051; transit access, B = 0.160). However, when county fixed effects were included, these relationships reversed. In the fully adjusted model with fixed effects, road network density shifted to a slightly negative association (B = -0.013), street intersection density became significantly negative (B = -0.065), and transit access shifted to a significantly negative association (B = -0.154). The unweighted results with fixed effects showed similar reversals (road network density, B = -0.007; intersection density, B = -0.060; transit access, B = -0.178). These reversals indicate reflect national-level patterns where non-Hispanic Black people/tracts may be located in counties with better overall walkability infrastructure.

Other variables for the non-Hispanic Black people/tracts showed less dramatic sensitivity. Land use mix remained consistently negative, dropping from -0.545 (unweighted, no fixed effects) to -0.532 (unweighted, with fixed effects) and from -0.542 to -0.534 (weighted). Fitness centers also remained negative, shifting from -0.388 to -0.458 (unweighted) and -0.400 to -0.515 (weighted), same as natural environment indicators and census self-response rate. For Hispanic people/tracts, road network density decreased from 0.302 to 0.070 (unweighted) and 0.317 to 0.084 (weighted) with fixed effects, while natural environment, such as NDVI shifts from -1.137 to -0.192 (unweighted) and -1.133 to -0.198 (weighted) and census self-response rate moves from -0.522 to -0.426 (unweighted) and -0.517 to -0.440 (weighted).

For high poverty people/tracts (relative to low poverty, <10%), road network density increased from 0.084 to 0.293 (unweighted) and 0.094 to 0.292 (weighted) with fixed effects, showing sensitivity. Census self-response rates were consistently negative, moving from -1.087 to -1.005 (unweighted) and -1.075 to -0.972 (weighted), with moderate sensitivity, while the relationship with some natural environment, including tree canopy cover, NDVI, and PM2.5 reversed, shifting from positive to negative when including fixed effects.

##### 1.3.2 With and Without Population Weighting

To interpret disparities from complementary perspectives, we estimated both unweighted and population-weighted linear regression models. In Model 1, population weighting introduced minor adjustments to the magnitude of coefficients, with little impact on direction or significance. For non-Hispanic Black people/tracts, without fixed effects, land use mix was nearly identical (-0.545 unweighted vs. -0.542 weighted), and fitness centers shifted slightly from -0.388 to -0.400. With fixed effects, land use mix remained stable (-0.532 unweighted vs. -0.534 weighted), and fitness centers moved from -0.458 to -0.515, reflecting modest refinements of 5-15%. Natural environment and social environment also showed minimal change.

For Hispanic people/tracts, road network density adjusted from 0.302 to 0.317 (no fixed effects) and 0.070 to 0.084 (with fixed effects), while NDVI shifted from -1.137 to -1.133 (no fixed effects) and -0.192 to -0.198 (with fixed effects), showing minor refinements, same as other dimension variables. For high-poverty people/tracts, fitness centers moved from -0.192 to -0.256 (no fixed effects) and -0.092 to -0.147 (with fixed effects), with changes typically within 5-15%. Census self-response rate adjusts from -1.087 to -1.075 (no fixed effects) and -1.005 to -0.972 (with fixed effects).

In summary, results were highly sensitive to the inclusion of county fixed effects, which can reverse the direction of built environment associations (e.g., road network density, intersection density, transit access for majority non-Hispanic Black tracts) and reduce the magnitude of others in general, distinguishing national place-based advantages from within-county inequities. In contrast, population weighting had a minimal effect, adjusting coefficients by 5-15% without changing their direction or significance, and provided complementary insights into place-based versus population-based disparities. Core disparities, such as lower fitness centers access, lower green space cover, and lower census self-response rate in non-Hispanic Black and high poverty people/tracts, remained robust across specifications, while built environment effects depend heavily on controlling for county-level variation.

#### 2. Supplementary Figures and Tables (Results)

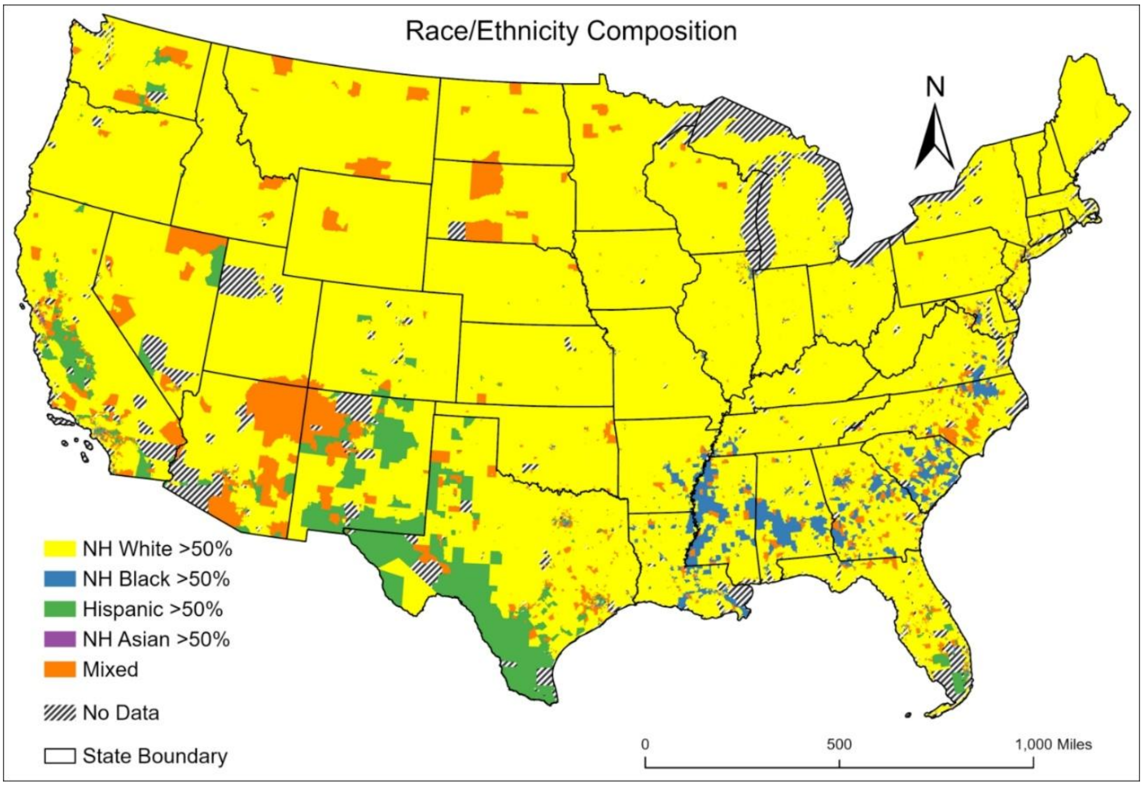

**Figure A1.** Spatial Distribution of Racial/Ethnic Composition across Census Tracts, Contiguous United States, 2018

**Notes:** Due to the limitations of visualizing all 69,889 tracts at a national scale, this map is intended to highlight broad spatial patterns rather than precise local variations.

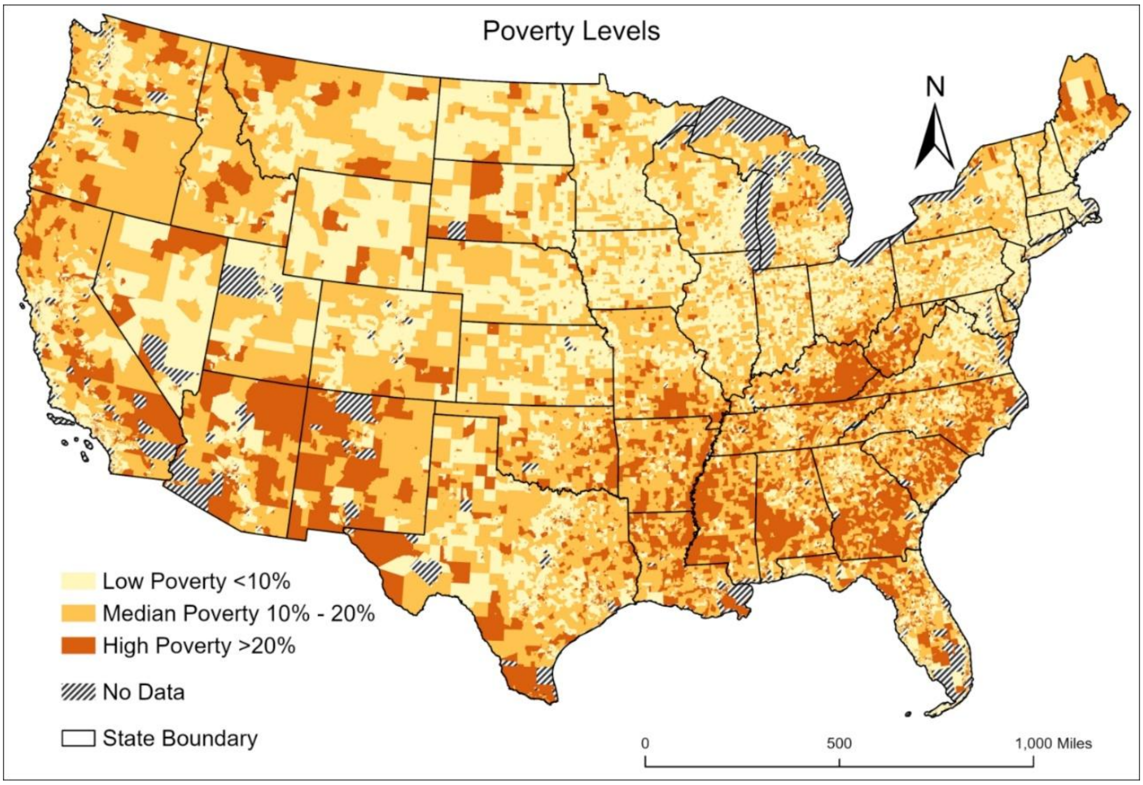

**Figure A2.** Spatial Distribution of Neighborhood Poverty Levels across Census Tracts, Contiguous United States, 2018

**Notes:** Due to the limitations of visualizing all 69,889 tracts at a national scale, this map is intended to highlight broad spatial patterns rather than precise local variations.

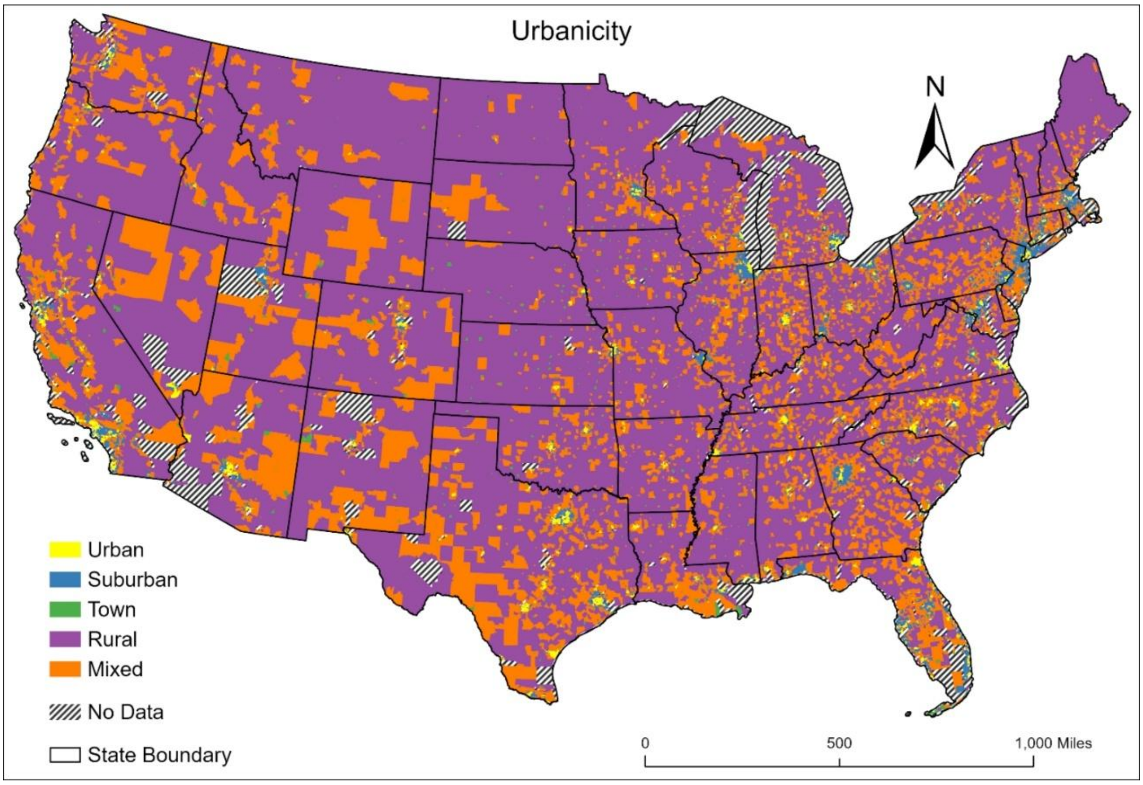

**Figure A3.** Spatial Distribution of Urbanicity Classifications across Census Tracts, Contiguous United States, 2010
**Notes:** Due to the limitations of visualizing all 69,889 tracts at a national scale, this map is intended to highlight broad spatial patterns rather than precise local variations.

**Table A1.** Number of Census Tracts by Racial/Ethnic Composition and Poverty Level Across Urbanicity Categories, Contiguous United States, 2018

| | **Total** | **W_NH >50%** | **B_NH >50%** | **Hisp >50%** | **A_NH >50%** | **Mixed >50%** | **LowPov** | **MedPov** | **HighPov** |
|---|---|---|---|---|---|---|---|---|---|
| National | 69889 | 48247 | 5753 | 6594 | 551 | 8744 | 29220 | 22598 | 18071 |
| Urban | 21856 | 10460 | 3551 | 3324 | 339 | 4182 | 6421 | 6367 | 9068 |
| Suburban | 21361 | 14542 | 1241 | 2234 | 198 | 3146 | 12315 | 5856 | 3190 |
| Town | 3446 | 2723 | 206 | 311 | 0 | 206 | 627 | 1321 | 1498 |
| Rural | 9754 | 9065 | 244 | 136 | 0 | 309 | 3472 | 4475 | 1807 |
| Mixed | 13472 | 11457 | 511 | 589 | 14 | 901 | 6385 | 4579 | 2508 |

**Notes:** Race/ethnicity categories include: Non-Hispanic White (W_NH), non-Hispanic Black (B_NH), Hispanic (Hisp), non-Hispanic Asian (A_NH), and Mixed (no majority racial or ethnic group). Poverty categories are defined as: Low poverty (<10%) (LowPov), medium poverty (10–20%) (MedPov), and high poverty (>20%) (HighPov).

**Table A2.** Associations Between Neighborhood Sociodemographic Composition and Physical Activity Environment Indicators, Contiguous U.S. Census Tracts, 2018

| | **Model 1: Race/Ethnicity** | | | **Model 2: Poverty** | |
|---|---|---|---|---|---|
| | B_NH >50% | Hisp >50% | A_NH >50% | MedPov | HighPov |
| LndMix | -0.421** | -0.192** | 0.048 | 0.034** | 0.055** |
| RoadD | 0.320** | 0.397** | 0.202** | 0.267** | 0.562** |
| IntD | 0.288** | 0.349** | 0.108** | 0.272** | 0.586** |
| TrnAcc | -0.003 | 0.136** | -0.046 | 0.111** | 0.242** |
| FitCtr | -0.532** | -0.640** | -0.388** | -0.122** | -0.276** |
| MemClb | -0.445** | -0.466** | -0.283** | -0.159** | -0.271** |
| RecCls | -0.474** | -0.490** | -0.157** | -0.099** | -0.218** |
| PubGlf | -0.222** | -0.276** | -0.055 | -0.183** | -0.312** |
| PrkCov | -0.148** | -0.339** | -0.143** | -0.115** | -0.192** |
| GrnSpc | -0.314** | -0.559** | -0.305** | -0.255** | -0.486** |
| TreCov | -0.252** | -0.358** | -0.252** | -0.152** | -0.336** |
| NDVIsm | -0.161** | -0.395** | -0.260** | -0.189** | -0.390** |
| PM25 | 0.336** | 0.535** | 0.342** | 0.186** | 0.379** |
| CnsRsp | -0.997** | -0.974** | -0.303** | -0.545** | -1.158** |

**Note:** Estimates are derived from multivariable linear regressions: Model 1 includes only race/ethnicity; Model 2 includes only poverty. All dependent variables are standardized (mean = 0, SD = 1). All regression models are weighted by population and include county fixed effects. Statistical significance is denoted as follows: ** $p < 0.05$; * $p < 0.10$. Reference categories are majority non-Hispanic White (race) and low poverty (<10%) (poverty). Race/ethnicity categories include: Non-Hispanic White (W_NH), non-Hispanic Black (B_NH), Hispanic (Hisp), non-Hispanic Asian (A_NH), and Mixed (no majority racial or ethnic group). Poverty categories are defined as: Low poverty (<10%) (LowPov), medium poverty (10–20%) (MedPov), and high poverty (>20%) (HighPov). All variables are from 2018 or the closest available year.

**Table A3.** Associations Between Neighborhood Sociodemographic Composition and Physical Activity Environment Indicators Across Model Specifications, Contiguous U.S. Census Tracts, 2018

| | **Model 1: Race/Ethnicity and Poverty with Urbanicity** | | | | | **Model 2: Race/Ethnicity** | | | **Model 3: Poverty** | |
|---|---|---|---|---|---|---|---|---|---|---|
| | B_NH >50% | Hisp >50% | A_NH >50% | MedPov | HighPov | B_NH >50% | Hisp >50% | A_NH >50% | MedPov | HighPov |
| **1) Unweighted with county fixed effects** | | | | | | | | | | |
| LndMix | -0.532** | -0.276** | -0.015 | 0.078** | 0.164** | -0.408** | -0.149** | 0.055 | 0.043** | 0.074** |
| RoadD | -0.007 | 0.070** | 0.014 | 0.154** | 0.293** | 0.348** | 0.392** | 0.194** | 0.268** | 0.584** |
| IntD | -0.060** | -0.010 | -0.074** | 0.182** | 0.366** | 0.319** | 0.338** | 0.113** | 0.277** | 0.607** |
| TrnAcc | -0.178** | -0.026* | -0.077** | 0.118** | 0.252** | -0.002 | 0.142** | 0.001** | 0.128** | 0.265** |
| FitCtr | -0.458** | -0.537** | -0.378** | -0.024** | -0.092** | -0.451** | -0.541** | -0.372** | -0.070** | -0.204** |
| MemClb | -0.349** | -0.350** | -0.205** | -0.064** | -0.083** | -0.374** | -0.388** | -0.225** | -0.112** | -0.204** |
| RecCls | -0.420** | -0.429** | -0.142** | -0.017* | -0.049** | -0.406** | -0.423** | -0.129** | -0.058** | -0.157** |
| PubGlf | -0.054** | -0.087** | 0.024 | -0.104** | -0.174** | -0.180** | -0.220** | -0.050 | -0.142** | -0.262** |
| PrkCov | -0.096** | -0.281** | -0.127** | -0.046** | -0.050** | -0.161** | -0.348** | -0.172** | -0.103** | -0.192** |
| GrnSpc | -0.077** | -0.306** | -0.175** | -0.151** | -0.239** | -0.333** | -0.548** | -0.312** | -0.259** | -0.506** |
| TreCov | -0.092** | -0.191** | -0.159** | -0.103** | -0.208** | -0.268** | -0.365** | -0.256** | -0.169** | -0.372** |
| NDVIsm | 0.052** | -0.192** | -0.151** | -0.135** | -0.263** | -0.171** | -0.408** | -0.269** | -0.202** | -0.419** |
| PM25 | 0.175** | 0.376** | 0.267** | 0.103** | 0.169** | 0.336** | 0.533** | 0.358** | 0.184** | 0.380** |
| CnsRsp | -0.555** | -0.426** | -0.059** | -0.470** | -1.005** | -1.079** | -0.977** | -0.314** | -0.552** | -1.207** |
| **2) Population-weighted without county fixed effects** | | | | | | | | | | |
| LndMix | -0.542** | -0.330** | -0.020 | 0.063** | 0.115** | -0.487** | -0.260** | -0.027 | -0.007 | -0.059** |
| RoadD | 0.141** | 0.317** | 0.614** | 0.059** | 0.094** | 0.690** | 0.858** | 1.292** | 0.041** | 0.406** |
| IntD | 0.039** | 0.212** | 0.515** | 0.085** | 0.172** | 0.548** | 0.705** | 1.083** | 0.074** | 0.424** |
| TrnAcc | 0.149** | 0.091** | 0.629** | 0.027** | 0.064** | 0.391** | 0.298** | 0.893** | 0.058** | 0.242** |
| FitCtr | -0.400** | -0.360** | -0.248** | -0.127** | -0.256** | -0.386** | -0.320** | -0.064 | -0.215** | -0.372** |
| MemClb | -0.288** | -0.269** | -0.177** | -0.136** | -0.213** | -0.351** | -0.319** | -0.128** | -0.196** | -0.329** |
| RecCls | -0.415** | -0.280** | 0.044 | -0.125** | -0.223** | -0.401** | -0.241** | 0.216** | -0.211** | -0.346** |
| PubGlf | -0.151** | -0.167** | -0.110** | -0.149** | -0.236** | -0.358** | -0.367** | -0.247** | -0.165** | -0.344** |
| PrkCov | -0.125** | -0.163** | 0.026 | -0.096** | -0.160** | -0.232** | -0.274** | -0.020 | -0.100** | -0.224** |
| GrnSpc | -0.119** | -0.443** | -0.508** | -0.053** | -0.068** | -0.501** | -0.824** | -0.975** | -0.065** | -0.350** |
| TreCov | 0.084** | -0.828** | -0.639** | 0.006 | 0.055** | -0.146** | -1.061** | -0.960** | -0.072** | -0.295** |
| NDVIsm | 0.079** | -1.133** | -1.157** | -0.035** | 0.011 | -0.246** | -1.452** | -1.591** | -0.124** | -0.420** |

| | | | | | | | | | | |
|---|---|---|---|---|---|---|---|---|---|---|
| PM25 | 0.499** | 0.488** | 0.778** | 0.051** | 0.140** | 0.755** | 0.742** | 1.036** | 0.072** | 0.394** |
| CnsRsp | -0.585** | -0.517** | -0.122** | -0.556** | -1.075** | -1.007** | -0.847** | 0.026 | -0.722** | -1.365** |
| **3) Unweighted without county fixed effects** | | | | | | | | | | |
| LndMix | -0.545** | -0.296** | -0.048 | 0.062** | 0.121** | -0.477** | -0.216** | -0.042 | -0.001 | -0.049** |
| RoadD | 0.151** | 0.302** | 0.583** | 0.058** | 0.084** | 0.773** | 0.884** | 1.308** | 0.015* | 0.420** |
| IntD | 0.051** | 0.191** | 0.466** | 0.088** | 0.171** | 0.629** | 0.722** | 1.074** | 0.057** | 0.442** |
| TrnAcc | 0.160** | 0.129** | 0.721** | 0.029** | 0.024** | 0.418** | 0.337** | 0.997** | 0.057** | 0.229** |
| FitCtr | -0.388** | -0.315** | -0.256** | -0.078** | -0.192** | -0.344** | -0.246** | -0.074* | -0.160** | -0.296** |
| MemClb | -0.295** | -0.246** | -0.157** | -0.096** | -0.159** | -0.326** | -0.263** | -0.100** | -0.150** | -0.267** |
| RecCls | -0.397** | -0.255** | 0.037 | -0.085** | -0.164** | -0.354** | -0.190** | 0.207** | -0.167** | -0.279** |
| PubGlf | -0.125** | -0.131** | -0.095** | -0.115** | -0.199** | -0.313** | -0.304** | -0.220** | -0.123** | -0.291** |
| PrkCov | -0.124** | -0.151** | 0.009 | -0.089** | -0.165** | -0.259** | -0.287** | -0.069 | -0.083** | -0.236** |
| GrnSpc | -0.160** | -0.415** | -0.489** | -0.055** | -0.054** | -0.588** | -0.822** | -0.991** | -0.047** | -0.350** |
| TreCov | 0.034** | -0.803** | -0.618** | -0.012 | 0.051** | -0.233** | -1.052** | -0.954** | -0.073** | -0.287** |
| NDVIsm | 0.066** | -1.137** | -1.147** | -0.043** | 0.017** | -0.294** | -1.472** | -1.596** | -0.106** | -0.389** |
| PM25 | 0.541** | 0.454** | 0.749** | 0.068** | 0.160** | 0.859** | 0.759** | 1.060** | 0.064** | 0.433** |
| CnsRsp | -0.632** | -0.522** | -0.135** | -0.559** | -1.087** | -1.066** | -0.825** | 0.036 | -0.737** | -1.396** |

**Notes:** This table compares estimates derived from multivariable linear regressions across three regression specifications: 1) Unweighted with county fixed effects; 2) Population-weighted without county fixed effects; 3) Unweighted without county fixed effects. Each specification includes one of three covariate combinations: Model 1 includes both race/ethnicity and poverty, with urbanicity controls; Model 2 includes only race/ethnicity; Model 3 includes only poverty. All dependent variables are standardized (mean = 0, SD = 1). All regression models are weighted by population and include county fixed effects. Statistical significance is denoted as follows: ** $p < 0.05$; * $p < 0.10$. Reference categories are majority non-Hispanic White (race) and low poverty (<10%) (poverty). Race/ethnicity categories include: Non-Hispanic White (W_NH), non-Hispanic Black (B_NH), Hispanic (Hisp), non-Hispanic Asian (A_NH), and Mixed (no majority racial or ethnic group). Poverty categories are defined as: Low poverty (<10%) (LowPov), medium poverty (10–20%) (MedPov), and high poverty (>20%) (HighPov).

**Table A4.** Associations Between Neighborhood Sociodemographic Composition and Physical Activity Environment Indicators, by Urbanicity, Contiguous U.S. Census Tracts, 2018 (Population-Weighted with County Fixed Effects)

| | **Model 1: Race/Ethnicity and Poverty with Urbanicity** | | | | | **Model 2: Race/Ethnicity** | | | **Model 3: Poverty** | |
|---|---|---|---|---|---|---|---|---|---|---|
| | B_NH >50% | Hisp >50% | A_NH >50% | MedPov | HighPov | B_NH >50% | Hisp >50% | A_NH >50% | MedPov | HighPov |
| **1) Urban (N = 21856 tracts)** | | | | | | | | | | |
| LndMix | -0.630** | -0.397** | -0.043 | 0.098** | 0.154** | -0.558** | -0.311** | 0.000 | 0.006 | -0.073** |
| RoadD | -0.198** | -0.091** | -0.166** | 0.205** | 0.424** | 0.008 | 0.146** | -0.047 | 0.176** | 0.355** |
| IntD | -0.224** | -0.195** | -0.263** | 0.187** | 0.429** | -0.013 | 0.046** | -0.143** | 0.140** | 0.325** |
| TrnAcc | -0.226** | -0.109** | -0.101** | 0.150** | 0.297** | -0.082** | 0.056** | -0.019 | 0.122** | 0.224** |
| FitCtr | -0.539** | -0.714** | -0.337** | 0.030 | -0.030 | -0.560** | -0.733** | -0.346** | -0.097** | -0.322** |
| MemClb | -0.425** | -0.464** | -0.284** | 0.031 | 0.023 | -0.416** | -0.452** | -0.279** | -0.057** | -0.182** |
| RecCls | -0.514** | -0.603** | -0.157** | 0.002 | -0.005 | -0.516** | -0.605** | -0.158** | -0.109** | -0.263** |
| PubGlf | -0.130** | -0.155** | 0.058 | -0.081** | -0.143** | -0.198** | -0.235** | 0.018 | -0.111** | -0.211** |
| PrkCov | -0.066** | -0.174** | -0.040 | -0.183** | -0.228** | -0.167** | -0.299** | -0.102* | -0.214** | -0.291** |
| GrnSpc | 0.024 | -0.136** | 0.043 | -0.281** | -0.480** | -0.204** | -0.402** | -0.090** | -0.304** | -0.518** |
| TreCov | 0.014 | -0.175** | -0.131** | -0.176** | -0.325** | -0.142** | -0.356** | -0.221** | -0.208** | -0.380** |
| NDVIsm | 0.185** | -0.147** | -0.101** | -0.171** | -0.322** | 0.030** | -0.326** | -0.191** | -0.187** | -0.333** |
| PM25 | 0.132** | 0.283** | 0.074** | 0.138** | 0.235** | 0.243** | 0.413** | 0.139** | 0.180** | 0.330** |
| CnsRsp | -0.429** | -0.404** | -0.065* | -0.465** | -1.058** | -0.951** | -0.997** | -0.361** | -0.557** | -1.262** |
| **2) Suburban (N = 21361 tracts)** | | | | | | | | | | |
| LndMix | -0.576** | -0.271** | -0.020 | 0.085** | 0.128** | -0.514** | -0.199** | 0.012 | 0.024 | -0.009 |
| RoadD | 0.083** | 0.260** | 0.170** | 0.312** | 0.479** | 0.310** | 0.526** | 0.290** | 0.362** | 0.565** |
| IntD | 0.022 | 0.178** | 0.087 | 0.314** | 0.514** | 0.263** | 0.457** | 0.213** | 0.348** | 0.571** |
| TrnAcc | -0.073** | 0.079** | -0.164** | 0.105** | 0.255** | 0.038 | 0.206** | -0.110* | 0.113** | 0.267** |
| FitCtr | -0.502** | -0.456** | -0.400** | -0.052** | -0.190** | -0.581** | -0.544** | -0.436** | -0.131** | -0.363** |
| MemClb | -0.381** | -0.322** | -0.123 | -0.108** | -0.180** | -0.465** | -0.419** | -0.167** | -0.171** | -0.310** |
| RecCls | -0.447** | -0.310** | -0.156* | -0.042** | -0.165** | -0.514** | -0.385** | -0.186** | -0.101** | -0.295** |
| PubGlf | -0.115** | -0.184** | 0.057 | -0.142** | -0.214** | -0.217** | -0.303** | 0.002 | -0.173** | -0.276** |
| PrkCov | -0.136** | -0.331** | -0.156** | -0.164** | -0.195** | -0.234** | -0.448** | -0.211** | -0.211** | -0.293** |
| GrnSpc | -0.145** | -0.502** | -0.319** | -0.307** | -0.435** | -0.355** | -0.748** | -0.432** | -0.388** | -0.588** |
| TreCov | -0.139** | -0.307** | -0.255** | -0.215** | -0.328** | -0.295** | -0.489** | -0.337** | -0.277** | -0.435** |
| NDVIsm | -0.025 | -0.330** | -0.219** | -0.196** | -0.323** | -0.176** | -0.505** | -0.299** | -0.249** | -0.417** |

| | | | | | | | | | | |
|---|---|---|---|---|---|---|---|---|---|---|
| PM25 | 0.175** | 0.421** | 0.302** | 0.142** | 0.216** | 0.278** | 0.541** | 0.356** | 0.203** | 0.342** |
| CnsRsp | -0.685** | -0.600** | -0.172** | -0.588** | -1.202** | -1.223** | -1.219** | -0.443** | -0.722** | -1.456** |
| **3) Town (N = 3446 tracts)** | | | | | | | | | | |
| LndMix | -0.222* | 0.372** | | 0.229** | 0.368** | -0.128 | 0.496** | | 0.253** | 0.404** |
| RoadD | 0.233* | -0.037 | | 0.269** | 0.450** | 0.352** | 0.115 | | 0.267** | 0.457** |
| IntD | 0.222* | 0.015 | | 0.270** | 0.492** | 0.360** | 0.183* | | 0.271** | 0.505** |
| TrnAcc | -0.027 | -0.055 | | 0.108** | 0.140** | 0.001 | -0.010 | | 0.105** | 0.133** |
| FitCtr | -0.436** | -0.309** | | 0.225** | 0.237** | -0.402** | -0.236* | | 0.214** | 0.196** |
| MemClb | -0.345** | 0.002 | | 0.052 | 0.068 | -0.331** | 0.024 | | 0.053 | 0.052 |
| RecCls | -0.204 | -0.060 | | 0.178** | 0.278** | -0.135 | 0.033 | | 0.177** | 0.268** |
| PubGlf | -0.162 | -0.251* | | -0.120* | -0.101 | -0.168 | -0.280** | | -0.134** | -0.136** |
| PrkCov | -0.177* | -0.291** | | -0.012 | 0.029 | -0.159* | -0.279** | | -0.030 | -0.014 |
| GrnSpc | -0.139 | -0.492** | | -0.140** | -0.281** | -0.223** | -0.589** | | -0.166** | -0.336** |
| TreCov | -0.084** | -0.100** | | -0.100** | -0.192** | -0.140** | -0.166** | | -0.107** | -0.209** |
| NDVIsm | 0.012 | 0.136** | | -0.142** | -0.267** | -0.064 | 0.044 | | -0.138** | -0.260** |
| PM25 | 0.162** | 0.704** | | 0.060** | 0.125** | 0.200** | 0.747** | | 0.093** | 0.193** |
| CnsRsp | -0.622** | -0.528** | | -0.397** | -0.978** | -0.946** | -0.876** | | -0.421** | -1.055** |
| **4) Rural (N = 9754 tracts)** | | | | | | | | | | |
| LndMix | 0.177* | 0.176 | | 0.067** | 0.147** | 0.213** | 0.219 | | 0.070** | 0.166** |
| RoadD | -0.137* | 0.092 | | -0.033 | 0.001 | -0.127 | 0.094 | | -0.036 | -0.023 |
| IntD | -0.030 | -0.074 | | 0.030 | 0.096** | -0.002 | -0.045 | | 0.027 | 0.081** |
| TrnAcc | -0.003 | -0.007 | | 0.023 | 0.018 | -0.003 | -0.003 | | 0.022 | 0.016 |
| FitCtr | 0.055 | -0.252 | | -0.094** | -0.096** | 0.046 | -0.279* | | -0.098** | -0.110** |
| MemClb | 0.065 | -0.200 | | -0.101** | -0.215** | 0.013 | -0.262 | | -0.105** | -0.232** |
| RecCls | -0.065 | -0.042 | | -0.064** | -0.103** | -0.085 | -0.071 | | -0.066** | -0.115** |
| PubGlf | -0.202* | -0.056 | | -0.017 | -0.089* | -0.231** | -0.083 | | -0.019 | -0.109** |
| PrkCov | -0.053 | -0.739** | | 0.052** | 0.038 | -0.054 | -0.729** | | 0.043** | -0.005 |
| GrnSpc | -0.228** | -0.611** | | 0.029** | 0.021 | -0.228** | -0.605** | | 0.023* | -0.003 |
| TreCov | -0.100** | -0.207** | | 0.040** | 0.035** | -0.098** | -0.197** | | 0.038** | 0.025** |
| NDVIsm | 0.053* | 0.051 | | 0.016* | -0.006 | 0.046 | 0.048 | | 0.016* | -0.005 |
| PM25 | 0.124** | 0.675** | | 0.044** | 0.099** | 0.149** | 0.704** | | 0.050** | 0.123** |
| CnsRsp | -0.248** | -0.224** | | -0.241** | -0.406** | -0.332** | -0.340** | | -0.248** | -0.453** |

| | | | | | | | | | | |
|---|---|---|---|---|---|---|---|---|---|---|
| **5) Mixed (N = 13472 tracts)** | | | | | | | | | | |
| LndMix | -0.272** | -0.111* | 0.131 | 0.145** | 0.281** | -0.149** | 0.031 | 0.176 | 0.139** | 0.251** |
| RoadD | 0.307** | 0.389** | 0.737** | 0.132** | 0.344** | 0.460** | 0.561** | 0.786** | 0.169** | 0.453** |
| IntD | 0.291** | 0.355** | 0.686** | 0.159** | 0.383** | 0.461** | 0.547** | 0.742** | 0.195** | 0.488** |
| TrnAcc | 0.152** | 0.063 | -0.431* | 0.095** | 0.146** | 0.214** | 0.138** | -0.405 | 0.105** | 0.180** |
| FitCtr | -0.348** | -0.446** | -0.661** | -0.131** | -0.142** | -0.406** | -0.520** | -0.692** | -0.156** | -0.227** |
| MemClb | -0.443** | -0.530** | -0.355 | -0.172** | -0.206** | -0.528** | -0.638** | -0.397 | -0.210** | -0.328** |
| RecCls | -0.210** | -0.265** | 0.009 | -0.056** | -0.079** | -0.243** | -0.306** | -0.006 | -0.071** | -0.131** |
| PubGlf | -0.077 | -0.235** | -0.170 | -0.138** | -0.241** | -0.182** | -0.357** | -0.211 | -0.153** | -0.285** |
| PrkCov | -0.067 | -0.409** | -0.207 | -0.022 | -0.025 | -0.077 | -0.422** | -0.213 | -0.048** | -0.100** |
| GrnSpc | -0.153** | -0.592** | -0.588** | -0.050** | -0.074** | -0.185** | -0.630** | -0.602** | -0.090** | -0.189** |
| TreCov | -0.135** | -0.287** | -0.224* | 0.014** | -0.021** | -0.146** | -0.296** | -0.223* | -0.007 | -0.083** |
| NDVIsm | -0.085** | -0.204** | -0.347** | -0.019** | -0.112** | -0.136** | -0.258** | -0.359** | -0.036** | -0.160** |
| PM25 | 0.231** | 0.433** | 0.343** | 0.092** | 0.185** | 0.312** | 0.526** | 0.372** | 0.122** | 0.278** |
| CnsRsp | -0.361** | -0.439** | 0.094 | -0.408** | -0.800** | -0.710** | -0.843** | -0.034 | -0.447** | -0.919** |

**Notes:** This table presents results from multivariable linear regressions estimated separately for census tracts classified as urban, suburban, town, rural, or mixed. Three model specifications are reported: Model 1 includes both race/ethnicity and poverty; Model 2 includes only race/ethnicity; Model 3 includes only poverty. All dependent variables are standardized (mean = 0, SD = 1). All regression models are weighted by population and include county fixed effects. Statistical significance is denoted as follows: ** p < 0.05; * p < 0.10. Reference categories are majority non-Hispanic White (race) and low poverty (<10%) (poverty). Race/ethnicity categories include: Non-Hispanic White (W_NH), non-Hispanic Black (B_NH), Hispanic (Hisp), non-Hispanic Asian (A_NH), and Mixed (no majority racial or ethnic group). Poverty categories are defined as: Low poverty (<10%) (LowPov), medium poverty (10–20%) (MedPov), and high poverty (>20%) (HighPov).

**Table A5.** Associations Between Neighborhood Sociodemographic Composition and Physical Activity Environment Indicators, by Urbanicity, Contiguous U.S. Census Tracts, 2018 (Unweighted with County Fixed Effects)

| | **Model 1: Race/Ethnicity and Poverty with Urbanicity** | | | | | **Model 2: Race/Ethnicity** | | | **Model 3: Poverty** | |
|---|---|---|---|---|---|---|---|---|---|---|
| | B_NH >50% | Hisp >50% | A_NH >50% | MedPov | HighPov | B_NH >50% | Hisp >50% | A_NH >50% | MedPov | HighPov |
| **1) Urban (N = 21856 tracts)** | | | | | | | | | | |
| LndMix | -0.632** | -0.361** | -0.022 | 0.094** | 0.179** | -0.547** | -0.262** | 0.036 | 0.010 | -0.055** |
| RoadD | -0.184** | -0.102** | -0.156** | 0.192** | 0.419** | 0.020 | 0.133** | -0.021 | 0.160** | 0.342** |
| IntD | -0.206** | -0.203** | -0.236** | 0.178** | 0.419** | 0.000 | 0.032 | -0.100* | 0.130** | 0.310** |
| TrnAcc | -0.257** | -0.130** | -0.056 | 0.176** | 0.336** | -0.097** | 0.057** | 0.052* | 0.145** | 0.246** |
| FitCtr | -0.484** | -0.618** | -0.353** | 0.055** | 0.008 | -0.488** | -0.617** | -0.352** | -0.056** | -0.258** |
| MemClb | -0.405** | -0.408** | -0.246** | 0.053** | 0.062** | -0.379** | -0.375** | -0.226** | -0.025 | -0.133** |
| RecCls | -0.469** | -0.527** | -0.149** | 0.028 | 0.028 | -0.458** | -0.512** | -0.140** | -0.068** | -0.210** |
| PubGlf | -0.098** | -0.114** | 0.054 | -0.067** | -0.126** | -0.158** | -0.185** | 0.014 | -0.088** | -0.178** |
| PrkCov | -0.093** | -0.155** | -0.107* | -0.149** | -0.193** | -0.177** | -0.260** | -0.168** | -0.180** | -0.259** |
| GrnSpc | 0.015 | -0.113** | 0.000 | -0.274** | -0.491** | -0.217** | -0.385** | -0.158** | -0.292** | -0.522** |
| TreCov | 0.027* | -0.158** | -0.105** | -0.203** | -0.378** | -0.152** | -0.368** | -0.227** | -0.229** | -0.420** |
| NDVIsm | 0.202** | -0.152** | -0.093** | -0.189** | -0.357** | 0.032** | -0.350** | -0.208** | -0.202** | -0.355** |
| PM25 | 0.128** | 0.267** | 0.093** | 0.130** | 0.225** | 0.234** | 0.391** | 0.165** | 0.168** | 0.315** |
| CnsRsp | -0.478** | -0.415** | -0.054 | -0.483** | -1.104** | -1.020** | -1.035** | -0.411** | -0.576** | -1.331** |
| **2) Suburban (N = 21361 tracts)** | | | | | | | | | | |
| LndMix | -0.540** | -0.233** | -0.065 | 0.088** | 0.126** | -0.477** | -0.161** | -0.031 | 0.034** | -0.008 |
| RoadD | 0.092** | 0.232** | 0.125** | 0.316** | 0.495** | 0.333** | 0.508** | 0.253** | 0.361** | 0.576** |
| IntD | 0.036 | 0.145** | 0.036 | 0.324** | 0.526** | 0.291** | 0.436** | 0.170** | 0.354** | 0.577** |
| TrnAcc | -0.048 | 0.082** | -0.201** | 0.114** | 0.274** | 0.079** | 0.222** | -0.140** | 0.121** | 0.288** |
| FitCtr | -0.428** | -0.407** | -0.383** | -0.024 | -0.144** | -0.491** | -0.472** | -0.409** | -0.092** | -0.301** |
| MemClb | -0.320** | -0.305** | -0.125* | -0.087** | -0.139** | -0.387** | -0.382** | -0.161** | -0.143** | -0.263** |
| RecCls | -0.378** | -0.303** | -0.162** | -0.018 | -0.110** | -0.426** | -0.353** | -0.182** | -0.072** | -0.237** |
| PubGlf | -0.066* | -0.155** | 0.024 | -0.108** | -0.179** | -0.152** | -0.254** | -0.021 | -0.131** | -0.227** |
| PrkCov | -0.131** | -0.336** | -0.136* | -0.158** | -0.182** | -0.224** | -0.446** | -0.190** | -0.204** | -0.281** |
| GrnSpc | -0.141** | -0.459** | -0.287** | -0.329** | -0.479** | -0.377** | -0.731** | -0.415** | -0.402** | -0.620** |
| TreCov | -0.131** | -0.289** | -0.233** | -0.242** | -0.372** | -0.313** | -0.497** | -0.330** | -0.299** | -0.475** |
| NDVIsm | -0.018 | -0.318** | -0.202** | -0.208** | -0.345** | -0.185** | -0.508** | -0.289** | -0.256** | -0.433** |

| | | | | | | | | | | |
|---|---|---|---|---|---|---|---|---|---|---|
| PM25 | 0.173** | 0.430** | 0.293** | 0.142** | 0.214** | 0.278** | 0.551** | 0.350** | 0.201** | 0.341** |
| CnsRsp | -0.723** | -0.584** | -0.137** | -0.606** | -1.248** | -1.311** | -1.239** | -0.429** | -0.736** | -1.518** |
| **3) Town (N = 3446 tracts)** | | | | | | | | | | |
| LndMix | -0.261** | 0.493** | | 0.216** | 0.401** | -0.144 | 0.635** | | 0.246** | 0.440** |
| RoadD | 0.245** | -0.019 | | 0.310** | 0.477** | 0.363** | 0.144 | | 0.308** | 0.489** |
| IntD | 0.246* | 0.017 | | 0.321** | 0.533** | 0.389** | 0.202 | | 0.321** | 0.549** |
| TrnAcc | -0.027 | -0.050 | | 0.152** | 0.164** | -0.003 | 0.001 | | 0.149** | 0.157** |
| FitCtr | -0.441** | -0.259** | | 0.131** | 0.148** | -0.418** | -0.212* | | 0.124** | 0.110* |
| MemClb | -0.296** | -0.067 | | 0.085 | 0.126* | -0.266** | -0.024 | | 0.085 | 0.107* |
| RecCls | -0.213* | -0.032 | | 0.128** | 0.225** | -0.151 | 0.046 | | 0.130** | 0.214** |
| PubGlf | -0.155 | -0.201 | | -0.136** | -0.121* | -0.164 | -0.237* | | -0.146** | -0.151** |
| PrkCov | -0.141 | -0.207** | | -0.027 | 0.020 | -0.121 | -0.197** | | -0.040 | -0.015 |
| GrnSpc | -0.131 | -0.376** | | -0.156** | -0.307** | -0.224** | -0.485** | | -0.175** | -0.351** |
| TreCov | -0.105 | -0.094 | | -0.110** | -0.213** | -0.168** | -0.170** | | -0.117** | -0.232** |
| NDVIsm | 0.008 | 0.115* | | -0.152** | -0.290** | -0.079 | 0.011 | | -0.150** | -0.288** |
| PM25 | 0.152** | 0.552** | | 0.073** | 0.132** | 0.190** | 0.599** | | 0.098** | 0.185** |
| CnsRsp | -0.598** | -0.552** | | -0.382** | -0.970** | -0.934** | -0.909** | | -0.404** | -1.047** |
| **4) Rural (N = 9754 tracts)** | | | | | | | | | | |
| LndMix | 0.170* | 0.144 | | 0.053* | 0.161** | 0.217** | 0.192 | | 0.055** | 0.177** |
| RoadD | -0.121 | 0.120 | | -0.019 | 0.035 | -0.103 | 0.132 | | -0.022 | 0.009 |
| IntD | -0.037 | 0.047 | | 0.045 | 0.153** | 0.009 | 0.093 | | 0.043 | 0.136** |
| TrnAcc | -0.004 | -0.008 | | 0.019 | 0.014 | -0.004 | -0.005 | | 0.018 | 0.011 |
| FitCtr | 0.043 | -0.263* | | -0.067** | -0.072 | 0.034 | -0.282** | | -0.071** | -0.086* |
| MemClb | 0.037 | -0.146 | | -0.088** | -0.201** | -0.015 | -0.204 | | -0.091** | -0.213** |
| RecCls | -0.068 | 0.061 | | -0.061** | -0.079* | -0.082 | 0.039 | | -0.061** | -0.086 |
| PubGlf | -0.200* | 0.031 | | -0.009 | -0.069 | -0.224** | 0.010 | | -0.011 | -0.087* |
| PrkCov | -0.099 | -0.682** | | 0.043** | 0.004 | -0.109 | -0.683** | | 0.031 | -0.056* |
| GrnSpc | -0.214** | -0.570** | | 0.034** | 0.039** | -0.209** | -0.560** | | 0.028** | 0.015 |
| TreCov | -0.086** | -0.215** | | 0.042** | 0.037** | -0.084** | -0.205** | | 0.039** | 0.027 |
| NDVIsm | 0.051* | -0.015 | | 0.014 | -0.012 | 0.042 | -0.020 | | 0.014 | -0.013 |
| PM25 | 0.129** | 0.635** | | 0.043** | 0.098** | 0.154** | 0.663** | | 0.049** | 0.122** |
| CnsRsp | -0.233** | -0.222** | | -0.209** | -0.377** | -0.319** | -0.329** | | -0.217** | -0.430** |

| | | | | | | | | | | |
|---|---|---|---|---|---|---|---|---|---|---|
| **5) Mixed (N = 13472 tracts)** | | | | | | | | | | |
| LndMix | -0.180** | -0.080 | 0.270 | 0.155** | 0.293** | -0.050 | 0.069 | 0.317 | 0.153** | 0.276** |
| RoadD | 0.333** | 0.371** | 0.398* | 0.119** | 0.319** | 0.479** | 0.532** | 0.443** | 0.155** | 0.431** |
| IntD | 0.328** | 0.324** | 0.410* | 0.149** | 0.362** | 0.492** | 0.508** | 0.463** | 0.184** | 0.470** |
| TrnAcc | 0.199** | 0.125* | -0.633** | 0.084** | 0.135** | 0.257** | 0.194** | -0.610** | 0.099** | 0.185** |
| FitCtr | -0.252** | -0.354** | -0.331 | -0.078** | -0.081** | -0.283** | -0.396** | -0.348 | -0.095** | -0.140** |
| MemClb | -0.294** | -0.413** | -0.402 | -0.136** | -0.175** | -0.366** | -0.504** | -0.436 | -0.162** | -0.261** |
| RecCls | -0.154** | -0.202** | 0.397 | -0.021** | -0.024** | -0.164** | -0.215** | 0.392 | -0.032 | -0.063** |
| PubGlf | -0.055 | -0.163** | -0.183 | -0.110** | -0.199** | -0.142** | -0.264** | -0.215 | -0.119** | -0.227** |
| PrkCov | -0.078 | -0.479** | -0.184 | -0.005 | -0.023 | -0.089** | -0.491** | -0.187 | -0.034* | -0.106** |
| GrnSpc | -0.200** | -0.599** | -0.360** | -0.057** | -0.079** | -0.233** | -0.640** | -0.375** | -0.096** | -0.194** |
| TreCov | -0.186** | -0.276** | -0.321** | 0.010 | -0.026 | -0.200** | -0.288** | -0.322** | -0.011 | -0.092** |
| NDVIsm | -0.109** | -0.201** | -0.377** | -0.021** | -0.109** | -0.161** | -0.255** | -0.390** | -0.038** | -0.159** |
| PM25 | 0.248** | 0.445** | 0.347** | 0.093** | 0.181** | 0.328** | 0.537** | 0.376** | 0.124** | 0.275** |
| CnsRsp | -0.388** | -0.402** | 0.125 | -0.420** | -0.820** | -0.751** | -0.819** | -0.005 | -0.457** | -0.939** |

**Notes:** This table presents results from multivariable linear regressions estimated separately for census tracts classified as urban, suburban, town, rural, or mixed. Three model specifications are reported: Model 1 includes both race/ethnicity and poverty; Model 2 includes only race/ethnicity; Model 3 includes only poverty. All dependent variables are standardized (mean = 0, SD = 1). All regression models are unweighted by population and include county fixed effects. Statistical significance is denoted as follows: ** $p < 0.05$; * $p < 0.10$. Reference categories are majority non-Hispanic White (race) and low poverty (<10%) (poverty). Race/ethnicity categories include: Non-Hispanic White (W_NH), non-Hispanic Black (B_NH), Hispanic (Hisp), non-Hispanic Asian (A_NH), and Mixed (no majority racial or ethnic group). Poverty categories are defined as: Low poverty (<10%) (LowPov), medium poverty (10–20%) (MedPov), and high poverty (>20%) (HighPov).

**Table A6.** Associations Between Neighborhood Sociodemographic Composition and Physical Activity Environment Indicators, by Urbanicity, Contiguous U.S. Census Tracts, 2018 (Population-Weighted, No County Fixed Effects)

| | **Model 1: Race/Ethnicity and Poverty with Urbanicity** | | | | | **Model 2: Race/Ethnicity** | | | **Model 3: Poverty** | |
|---|---|---|---|---|---|---|---|---|---|---|
| | B_NH >50% | Hisp >50% | A_NH >50% | MedPov | HighPov | B_NH >50% | Hisp >50% | A_NH >50% | MedPov | HighPov |
| **1) Urban (N = 21856 tracts)** | | | | | | | | | | |
| LndMix | -0.703** | -0.398** | -0.087 | 0.091** | 0.158** | -0.627** | -0.333** | -0.091* | -0.010 | -0.103** |
| RoadD | 0.128** | 0.261** | 0.709** | 0.095** | 0.181** | 0.215** | 0.336** | 0.705** | 0.138** | 0.267** |
| IntD | -0.055** | 0.057** | 0.411** | 0.105** | 0.248** | 0.068** | 0.161** | 0.406** | 0.108** | 0.244** |
| TrnAcc | 0.184** | 0.087** | 0.701** | 0.071** | 0.094** | 0.226** | 0.125** | 0.698** | 0.095** | 0.150** |
| FitCtr | -0.502** | -0.426** | -0.219** | -0.061** | -0.205** | -0.607** | -0.512** | -0.216** | -0.147** | -0.420** |
| MemClb | -0.378** | -0.311** | -0.146** | -0.034** | -0.098** | -0.428** | -0.352** | -0.144** | -0.097** | -0.258** |
| RecCls | -0.516** | -0.379** | 0.037 | -0.065** | -0.151** | -0.591** | -0.442** | 0.040 | -0.148** | -0.363** |
| PubGlf | -0.203** | -0.196** | -0.046 | -0.088** | -0.151** | -0.275** | -0.258** | -0.042 | -0.130** | -0.248** |
| PrkCov | -0.094** | -0.203** | 0.065 | -0.224** | -0.288** | -0.224** | -0.320** | 0.073 | -0.259** | -0.362** |
| GrnSpc | -0.084** | -0.454** | -0.674** | -0.203** | -0.310** | -0.229** | -0.581** | -0.666** | -0.273** | -0.438** |
| TreCov | 0.136** | -0.821** | -0.812** | -0.095** | -0.115** | 0.085** | -0.867** | -0.809** | -0.202** | -0.283** |
| NDVIsm | 0.144** | -1.048** | -1.159** | -0.001 | 0.087** | 0.193** | -1.010** | -1.160** | -0.141** | -0.134** |
| PM25 | 0.697** | 0.607** | 0.693** | 0.064** | 0.166** | 0.780** | 0.677** | 0.690** | 0.188** | 0.468** |
| CnsRsp | -0.679** | -0.479** | -0.208** | -0.499** | -1.091** | -1.215** | -0.932** | -0.185** | -0.616** | -1.371** |
| **2) Suburban (N = 21361 tracts)** | | | | | | | | | | |
| LndMix | -0.545** | -0.344** | 0.017 | 0.067** | 0.103** | -0.502** | -0.294** | 0.024 | -0.024 | -0.090** |
| RoadD | 0.225** | 0.682** | 0.914** | 0.190** | 0.233** | 0.328** | 0.803** | 0.930** | 0.337** | 0.507** |
| IntD | 0.095** | 0.482** | 0.752** | 0.205** | 0.316** | 0.227** | 0.636** | 0.771** | 0.306** | 0.502** |
| TrnAcc | 0.069** | 0.150** | 0.081 | 0.006 | 0.089** | 0.099** | 0.184** | 0.084 | 0.037** | 0.151** |
| FitCtr | -0.296** | -0.303** | -0.332** | -0.112** | -0.294** | -0.407** | -0.431** | -0.345** | -0.171** | -0.429** |
| MemClb | -0.275** | -0.245** | -0.210** | -0.130** | -0.217** | -0.364** | -0.349** | -0.223** | -0.186** | -0.336** |
| RecCls | -0.371** | -0.202** | -0.069 | -0.116** | -0.292** | -0.482** | -0.330** | -0.083 | -0.162** | -0.403** |
| PubGlf | -0.186** | -0.252** | -0.128* | -0.119** | -0.172** | -0.259** | -0.337** | -0.139* | -0.175** | -0.285** |
| PrkCov | -0.077** | -0.216** | 0.049 | -0.240** | -0.323** | -0.216** | -0.379** | 0.028 | -0.271** | -0.401** |
| GrnSpc | 0.031 | -0.723** | -0.758** | -0.274** | -0.354** | -0.124** | -0.904** | -0.782** | -0.399** | -0.599** |
| TreCov | 0.210** | -0.946** | -0.886** | -0.246** | -0.339** | 0.065** | -1.116** | -0.908** | -0.407** | -0.641** |
| NDVIsm | 0.126** | -1.361** | -1.469** | -0.174** | -0.197** | 0.037 | -1.465** | -1.484** | -0.416** | -0.656** |

| | | | | | | | | | | |
|---|---|---|---|---|---|---|---|---|---|---|
| PM25 | 0.457** | 0.510** | 1.113** | 0.027* | 0.118** | 0.499** | 0.558** | 1.118** | 0.134** | 0.346** |
| CnsRsp | -0.660** | -0.589** | -0.008 | -0.644** | -1.309** | -1.177** | -1.186** | -0.077 | -0.799** | -1.612** |
| **3) Town (N = 3446 tracts)** | | | | | | | | | | |
| LndMix | -0.141* | -0.397** | | 0.295** | 0.311** | -0.064 | -0.337** | | 0.267** | 0.234** |
| RoadD | -0.108 | -0.165** | | 0.181** | 0.355** | 0.032 | -0.079 | | 0.168** | 0.316** |
| IntD | -0.132* | -0.165** | | 0.192** | 0.398** | 0.029 | -0.067 | | 0.180** | 0.358** |
| TrnAcc | -0.090 | -0.079 | | 0.045 | 0.028 | -0.090 | -0.076 | | 0.039 | 0.005 |
| FitCtr | -0.396** | -0.384** | | 0.029 | -0.027 | -0.421** | -0.396** | | 0.000 | -0.127** |
| MemClb | -0.106 | -0.273** | | -0.079 | -0.115** | -0.145* | -0.299** | | -0.100** | -0.174** |
| RecCls | -0.312** | -0.367** | | -0.064 | -0.066 | -0.327** | -0.380** | | -0.092* | -0.158** |
| PubGlf | -0.250** | -0.156** | | -0.148** | -0.205** | -0.316** | -0.201** | | -0.164** | -0.263** |
| PrkCov | -0.292** | -0.257** | | -0.109** | -0.215** | -0.377** | -0.310** | | -0.128** | -0.284** |
| GrnSpc | -0.002 | -0.168** | | -0.092* | -0.119** | -0.038 | -0.194** | | -0.098** | -0.129** |
| TreCov | 0.653** | -0.888** | | -0.002 | 0.134** | 0.730** | -0.848** | | -0.054 | 0.066 |
| NDVIsm | 0.393** | -1.326** | | 0.011 | 0.028 | 0.405** | -1.319** | | -0.081* | -0.163** |
| PM25 | 0.191** | 0.303** | | 0.130** | 0.406** | 0.377** | 0.410** | | 0.146** | 0.454** |
| CnsRsp | -0.638** | -0.647** | | -0.373** | -0.923** | -1.036** | -0.882** | | -0.434** | -1.126** |
| **4) Rural (N = 9754 tracts)** | | | | | | | | | | |
| LndMix | 0.096 | 0.523** | | -0.158** | -0.321** | -0.056 | 0.430** | | -0.151** | -0.289** |
| RoadD | -0.335** | -0.264** | | -0.116** | -0.134** | -0.387** | -0.302** | | -0.129** | -0.222** |
| IntD | -0.102** | -0.039 | | -0.032* | 0.003 | -0.094* | -0.038 | | -0.035** | -0.026 |
| TrnAcc | -0.007 | -0.014 | | -0.022 | -0.040 | -0.026 | -0.026 | | -0.022 | -0.042 |
| FitCtr | -0.064 | -0.129 | | -0.234** | -0.278** | -0.175** | -0.209** | | -0.238** | -0.308** |
| MemClb | 0.210** | -0.284** | | -0.151** | -0.317** | 0.058 | -0.376** | | -0.153** | -0.319** |
| RecCls | -0.119 | 0.057 | | -0.186** | -0.256** | -0.227** | -0.017 | | -0.188** | -0.277** |
| PubGlf | -0.128* | -0.050 | | -0.157** | -0.365** | -0.307** | -0.156 | | -0.161** | -0.392** |
| PrkCov | -0.317** | 0.156* | | 0.089** | 0.071** | -0.295** | 0.176** | | 0.085** | 0.033 |
| GrnSpc | -0.407** | -0.127 | | 0.298** | 0.521** | -0.169** | 0.023 | | 0.293** | 0.488** |
| TreCov | 0.301** | -1.454** | | 0.345** | 0.660** | 0.610** | -1.263** | | 0.327** | 0.585** |
| NDVIsm | 0.213** | -2.217** | | 0.002 | 0.036 | 0.234** | -2.206** | | -0.030 | -0.125** |
| PM25 | 0.291** | 0.053 | | 0.064** | 0.046 | 0.304** | 0.066 | | 0.058** | -0.010 |
| CnsRsp | -0.345** | -1.247** | | -0.582** | -0.960** | -0.776** | -1.524** | | -0.611** | -1.143** |

| | | | | | | | | | | |
|---|---|---|---|---|---|---|---|---|---|---|
| **5) Mixed (N = 13472 tracts)** | | | | | | | | | | |
| LndMix | -0.226** | -0.316** | -0.096 | 0.114** | 0.235** | -0.107** | -0.215** | -0.123 | 0.100** | 0.157** |
| RoadD | 0.396** | 0.640** | 2.024** | -0.113** | -0.108** | 0.348** | 0.595** | 2.037** | -0.063** | 0.116** |
| IntD | 0.379** | 0.599** | 2.176** | -0.068** | -0.047 | 0.360** | 0.580** | 2.182** | -0.019 | 0.170** |
| TrnAcc | 0.098* | 0.129** | -0.069 | -0.024** | -0.037** | 0.080* | 0.113** | -0.065 | -0.010 | 0.021 |
| FitCtr | -0.155** | -0.184** | -0.198 | -0.262** | -0.351** | -0.324** | -0.334** | -0.156 | -0.264** | -0.378** |
| MemClb | -0.152** | -0.247** | -0.334 | -0.239** | -0.354** | -0.325** | -0.398** | -0.292 | -0.250** | -0.412** |
| RecCls | -0.138** | -0.063 | 0.516** | -0.216** | -0.313** | -0.290** | -0.196** | 0.554** | -0.216** | -0.325** |
| PubGlf | -0.180** | -0.203** | -0.573** | -0.194** | -0.331** | -0.345** | -0.345** | -0.534** | -0.208** | -0.397** |
| PrkCov | -0.234** | -0.104** | -0.010 | -0.058** | -0.118** | -0.293** | -0.155** | 0.004 | -0.070** | -0.177** |
| GrnSpc | -0.294** | -0.452** | -1.090** | 0.061** | 0.069** | -0.261** | -0.423** | -1.098** | 0.030 | -0.073** |
| TreCov | 0.249** | -1.345** | -0.800** | 0.142** | 0.246** | 0.372** | -1.239** | -0.829** | 0.087** | 0.020 |
| NDVIsm | 0.044 | -1.801** | -1.519** | 0.109** | 0.167** | 0.127** | -1.730** | -1.539** | 0.024 | -0.194** |
| PM25 | 0.315** | 0.511** | 1.026** | 0.074** | 0.160** | 0.396** | 0.580** | 1.008** | 0.104** | 0.301** |
| CnsRsp | -0.380** | -0.701** | 0.423** | -0.694** | -1.181** | -0.968** | -1.209** | 0.562** | -0.738** | -1.384** |

**Notes:** This table presents results from multivariable linear regressions estimated separately for census tracts classified as urban, suburban, town, rural, or mixed. Three model specifications are reported: Model 1 includes both race/ethnicity and poverty; Model 2 includes only race/ethnicity; Model 3 includes only poverty. All dependent variables are standardized (mean = 0, SD = 1). All regression models are weighted by population and exclude county fixed effects. Statistical significance is denoted as follows: ** $p < 0.05$; * $p < 0.10$. Reference categories are majority non-Hispanic White (race) and low poverty (<10%) (poverty). Race/ethnicity categories include: Non-Hispanic White (W_NH), non-Hispanic Black (B_NH), Hispanic (Hisp), non-Hispanic Asian (A_NH), and Mixed (no majority racial or ethnic group). Poverty categories are defined as: Low poverty (<10%) (LowPov), medium poverty (10–20%) (MedPov), and high poverty (>20%) (HighPov).

**Table A7.** Associations Between Neighborhood Sociodemographic Composition and Physical Activity Environment Indicators, by Urbanicity, Contiguous U.S. Census Tracts, 2018 (Unweighted, No County Fixed Effects)

| | **Model 1: Race/Ethnicity and Poverty with Urbanicity** | | | | | **Model 2: Race/Ethnicity** | | | **Model 3: Poverty** | |
|---|---|---|---|---|---|---|---|---|---|---|
| | B_NH >50% | Hisp >50% | A_NH >50% | MedPov | HighPov | B_NH >50% | Hisp >50% | A_NH >50% | MedPov | HighPov |
| **1) Urban (N = 21856 tracts)** | | | | | | | | | | |
| LndMix | -0.723** | -0.394** | -0.112** | 0.095** | 0.201** | -0.625** | -0.312** | -0.112** | -0.002 | -0.084** |
| RoadD | 0.167** | 0.271** | 0.714** | 0.078** | 0.121** | 0.223** | 0.320** | 0.714** | 0.123** | 0.220** |
| IntD | -0.023 | 0.057** | 0.388** | 0.095** | 0.219** | 0.085** | 0.146** | 0.389** | 0.100** | 0.220** |
| TrnAcc | 0.174** | 0.125** | 0.803** | 0.079** | 0.038** | 0.183** | 0.138** | 0.803** | 0.111** | 0.111** |
| FitCtr | -0.460** | -0.372** | -0.254** | -0.031* | -0.156** | -0.543** | -0.437** | -0.255** | -0.104** | -0.358** |
| MemClb | -0.379** | -0.282** | -0.139** | -0.004 | -0.052** | -0.407** | -0.304** | -0.139** | -0.061** | -0.214** |
| RecCls | -0.476** | -0.329** | 0.025 | -0.035** | -0.112** | -0.534** | -0.376** | 0.025 | -0.105** | -0.313** |
| PubGlf | -0.175** | -0.157** | -0.045 | -0.073** | -0.123** | -0.233** | -0.207** | -0.046 | -0.106** | -0.207** |
| PrkCov | -0.103** | -0.176** | 0.010 | -0.200** | -0.277** | -0.227** | -0.286** | 0.008 | -0.228** | -0.344** |
| GrnSpc | -0.142** | -0.453** | -0.710** | -0.185** | -0.273** | -0.266** | -0.562** | -0.712** | -0.254** | -0.406** |
| TreCov | 0.066** | -0.798** | -0.799** | -0.110** | -0.121** | 0.016 | -0.844** | -0.800** | -0.205** | -0.267** |
| NDVIsm | 0.115** | -1.062** | -1.151** | -0.005 | 0.116** | 0.181** | -1.012** | -1.151** | -0.133** | -0.072** |
| PM25 | 0.712** | 0.568** | 0.669** | 0.072** | 0.174** | 0.799** | 0.640** | 0.670** | 0.186** | 0.484** |
| CnsRsp | -0.740** | -0.493** | -0.265** | -0.511** | -1.117** | -1.287** | -0.949** | -0.270 | -0.628** | -1.434** |
| **2) Suburban (N = 21361 tracts)** | | | | | | | | | | |
| LndMix | -0.521 | -0.318** | -0.037 | 0.077** | 0.113** | -0.472** | -0.264** | -0.032 | -0.002 | -0.066** |
| RoadD | 0.243 | 0.661** | 0.811** | 0.197** | 0.250** | 0.356** | 0.785** | 0.824** | 0.328** | 0.500** |
| IntD | 0.115 | 0.451** | 0.648** | 0.221** | 0.339** | 0.263** | 0.613** | 0.663** | 0.310** | 0.503** |
| TrnAcc | 0.156 | 0.194** | 0.010 | -0.013 | 0.072** | 0.182** | 0.218** | 0.011 | 0.029* | 0.159** |
| FitCtr | -0.289 | -0.254** | -0.282** | -0.080** | -0.249** | -0.390** | -0.358** | -0.289** | -0.127** | -0.363** |
| MemClb | -0.272 | -0.228** | -0.175** | -0.102** | -0.169** | -0.346** | -0.308** | -0.183** | -0.153** | -0.281** |
| RecCls | -0.337 | -0.189** | -0.056 | -0.084** | -0.227** | -0.430** | -0.286** | -0.063 | -0.125** | -0.331** |
| PubGlf | -0.148 | -0.212** | -0.129* | -0.084** | -0.139** | -0.209** | -0.277** | -0.135* | -0.126** | -0.228** |
| PrkCov | -0.033 | -0.200** | 0.059 | -0.249** | -0.343** | -0.186** | -0.368** | 0.042 | -0.273** | -0.401** |
| GrnSpc | -0.033 | -0.685** | -0.711** | -0.307** | -0.400** | -0.213** | -0.883** | -0.731** | -0.417** | -0.613** |
| TreCov | 0.165 | -0.913** | -0.860** | -0.294** | -0.385** | -0.008 | -1.103** | -0.880** | -0.432** | -0.639** |
| NDVIsm | 0.101 | -1.394** | -1.462** | -0.181** | -0.189** | 0.012 | -1.494** | -1.474** | -0.403** | -0.602** |

| | | | | | | | | | | |
|---|---|---|---|---|---|---|---|---|---|---|
| PM25 | 0.528 | 0.512** | 1.081** | 0.052** | 0.162** | 0.593** | 0.580** | 1.086** | 0.152** | 0.389** |
| CnsRsp | -0.669 | -0.550** | 0.044 | -0.676** | -1.373** | -1.249** | -1.167** | -0.008 | -0.812** | -1.658** |
| **3) Town (N = 3446 tracts)** | | | | | | | | | | |
| LndMix | -0.137* | -0.282** | | 0.276** | 0.307** | -0.061 | -0.224** | | 0.262** | 0.257** |
| RoadD | -0.105 | -0.165** | | 0.191** | 0.381** | 0.043 | -0.071 | | 0.182** | 0.345** |
| IntD | -0.121 | -0.175** | | 0.211** | 0.427** | 0.046 | -0.070 | | 0.201** | 0.388** |
| TrnAcc | -0.085 | -0.084 | | 0.055 | 0.000 | -0.105 | -0.092 | | 0.050 | -0.023 |
| FitCtr | -0.375** | -0.319** | | 0.006 | -0.055 | -0.408** | -0.337** | | -0.013 | -0.141** |
| MemClb | -0.081 | -0.239** | | -0.061 | -0.101 | -0.116 | -0.262** | | -0.073 | -0.144** |
| RecCls | -0.288** | -0.301** | | -0.058 | -0.065 | -0.304** | -0.313** | | -0.076 | -0.141** |
| PubGlf | -0.205** | -0.108* | | -0.170** | -0.231** | -0.276** | -0.157** | | -0.179** | -0.280** |
| PrkCov | -0.279** | -0.235** | | -0.175** | -0.277** | -0.374** | -0.298** | | -0.189** | -0.344** |
| GrnSpc | 0.049 | -0.018 | | -0.097** | -0.141** | 0.003 | -0.049 | | -0.093* | -0.122** |
| TreCov | 0.623** | -0.940** | | 0.016 | 0.124** | 0.687** | -0.903** | | -0.021 | 0.084* |
| NDVIsm | 0.359** | -1.366** | | 0.066 | 0.067 | 0.374** | -1.354** | | -0.001 | -0.088* |
| PM25 | 0.150** | 0.128** | | 0.205** | 0.485** | 0.351** | 0.253** | | 0.208** | 0.503** |
| CnsRsp | -0.660** | -0.715** | | -0.290** | -0.822** | -1.022** | -0.934** | | -0.341** | -1.035** |
| **4) Rural (N = 9754 tracts)** | | | | | | | | | | |
| LndMix | 0.039 | 0.565** | | -0.170** | -0.380** | -0.148** | 0.455** | | -0.163** | -0.346** |
| RoadD | -0.287** | -0.301** | | -0.123** | -0.127** | -0.334** | -0.335** | | -0.137** | -0.222** |
| IntD | -0.084 | -0.047 | | -0.035 | 0.020 | -0.064 | -0.040 | | -0.040* | -0.013 |
| TrnAcc | -0.007 | -0.015 | | -0.029 | -0.047 | -0.028 | -0.028 | | -0.030 | -0.049* |
| FitCtr | -0.041 | -0.137 | | -0.174** | -0.231** | -0.139** | -0.201** | | -0.178** | -0.258** |
| MemClb | 0.112* | -0.226** | | -0.128** | -0.294** | -0.034 | -0.312** | | -0.130** | -0.302** |
| RecCls | -0.106 | 0.072 | | -0.149** | -0.201** | -0.191** | 0.016 | | -0.151** | -0.221** |
| PubGlf | -0.139** | -0.027 | | -0.130** | -0.320** | -0.300** | -0.121 | | -0.133** | -0.345** |
| PrkCov | -0.375** | 0.117 | | 0.089** | 0.078** | -0.350** | 0.138 | | 0.083** | 0.024 |
| GrnSpc | -0.451** | -0.015 | | 0.298** | 0.565** | -0.184** | 0.147* | | 0.294** | 0.533** |
| TreCov | 0.306** | -1.445** | | 0.327** | 0.688** | 0.641** | -1.246** | | 0.309** | 0.609** |
| NDVIsm | 0.261** | -2.294** | | -0.008 | 0.029 | 0.280** | -2.285** | | -0.041* | -0.138** |
| PM25 | 0.361** | -0.002 | | 0.065** | 0.065** | 0.385** | 0.015 | | 0.058** | 0.012 |
| CnsRsp | -0.286** | -1.254** | | -0.542** | -0.908** | -0.702** | -1.513** | | -0.572** | -1.098** |

| | | | | | | | | | | |
|---|---|---|---|---|---|---|---|---|---|---|
| **5) Mixed (N = 13472 tracts)** | | | | | | | | | | |
| LndMix | -0.164** | -0.269** | 0.068 | 0.138** | 0.246** | -0.035 | -0.163** | 0.038 | 0.127** | 0.188** |
| RoadD | 0.392** | 0.611** | 1.970** | -0.110** | -0.087** | 0.356** | 0.575** | 1.979** | -0.063** | 0.126** |
| IntD | 0.379** | 0.572** | 2.241** | -0.062** | -0.020 | 0.376** | 0.565** | 2.243** | -0.016 | 0.187** |
| TrnAcc | 0.143** | 0.188** | -0.068 | -0.038** | -0.053** | 0.117** | 0.165** | -0.061 | -0.022 | 0.021 |
| FitCtr | -0.134** | -0.146** | -0.091 | -0.162** | -0.233** | -0.251** | -0.246** | -0.064 | -0.163** | -0.252** |
| MemClb | -0.128** | -0.234** | -0.396 | -0.182** | -0.286** | -0.274** | -0.356** | -0.362 | -0.192** | -0.335** |
| RecCls | -0.132** | -0.073* | 0.629** | -0.160** | -0.231** | -0.248** | -0.172** | 0.656** | -0.162** | -0.251** |
| PubGlf | -0.167** | -0.168** | -0.507* | -0.154** | -0.287** | -0.318** | -0.293** | -0.472* | -0.166** | -0.344** |
| PrkCov | -0.241** | -0.117** | 0.156 | -0.060** | -0.155** | -0.326** | -0.185** | 0.174 | -0.071** | -0.219** |
| GrnSpc | -0.371** | -0.438** | -0.779** | 0.041** | 0.065** | -0.338** | -0.410** | -0.786** | 0.012 | -0.080** |
| TreCov | 0.180** | -1.356** | -0.906** | 0.102** | 0.208** | 0.290** | -1.266** | -0.931** | 0.050** | 0.009 |
| NDVIsm | 0.021 | -1.838** | -1.826** | 0.087** | 0.142** | 0.094** | -1.776** | -1.843** | 0.005 | -0.184** |
| PM25 | 0.348** | 0.431** | 1.024** | 0.087** | 0.186** | 0.448** | 0.512** | 1.001** | 0.111** | 0.311** |
| CnsRsp | -0.395** | -0.693** | 0.570** | -0.705** | -1.193** | -1.012** | -1.209** | 0.713** | -0.747** | -1.390** |

**Notes:** This table presents results from multivariable linear regressions estimated separately for census tracts classified as urban, suburban, town, rural, or mixed. Three model specifications are reported: Model 1 includes both race/ethnicity and poverty; Model 2 includes only race/ethnicity; Model 3 includes only poverty. All dependent variables are standardized (mean = 0, SD = 1). All regression models are unweighted by population and exclude county fixed effects. Statistical significance is denoted as follows: ** $p < 0.05$; * $p < 0.10$. Reference categories are majority non-Hispanic White (race) and low poverty (<10%) (poverty). Race/ethnicity categories include: Non-Hispanic White (W_NH), non-Hispanic Black (B_NH), Hispanic (Hisp), non-Hispanic Asian (A_NH), and Mixed (no majority racial or ethnic group). Poverty categories are defined as: Low poverty (<10%) (LowPov), medium poverty (10–20%) (MedPov), and high poverty (>20%) (HighPov).
